# Predicting ICU Transfer and Short-term Mortality in Emergency Department Atrial Fibrillation Patients: An Enhanced Machine Learning Model Using MIMIC Data

**DOI:** 10.1101/2025.09.22.25336382

**Authors:** Tinghuan Li, Zhuoyang Li, Shuheng Chen, Elham Pishgar, Kamiar Alaei, Greg Placencia, Maryam Pishgar

## Abstract

Atrial fibrillation (AF) is a prevalent condition in emergency department (ED) patients and is associated with an elevated risk of intensive care unit (ICU) transfer and short-term mortality. Early identification of high-risk patients is critical for timely intervention and improved clinical outcomes. We constructed a combined cohort from the MIMIC-IV-ED and MIMIC-IV databases, comprising ED admissions of patients with AF, and developed an interpretable machine learning (ML) framework to predict ICU transfer and mortality within 3, 7, and 30 days using clinical variables obtained at triage. The preprocessing pipeline included imputation of missing data, z-score normalization of numerical features, one-hot encoding of categorical variables, and correction for class imbalance using the Synthetic Minority Over-sampling Technique (SMOTE). A hybrid feature selection strategy combining Recursive Feature Elimination with Cross-Validation (RFECV) and Least Absolute Shrinkage and Selection Operator (LASSO) regression reduced the initial set to 19 clinically relevant predictors. Among six evaluated machine learning algorithms, LightGBM demonstrated the highest performance for ICU transfer (AUROC = 0.7979, 95% confidence interval (CI): 0.7916–0.8041) and for 7- and 30-day mortality (AUROC = 0.8316, 95% CI: 0.8156–0.8476; AUROC = 0.8010, 95% CI: 0.7898–0.8123), while CatBoost achieved the best performance for 3-day mortality (AUROC = 0.8444, 95% CI: 0.8237–0.8644). SHAP(SHapley Additive exPlanations) analysis identified O_2_sat, acuity, and resprate as key determinants, underscoring the clinical plausibility and interpretability of the models. These findings highlight the potential of interpretable machine learning approaches to enable early, time-sensitive risk stratification and support informed clinical decision-making for AF patients in the ED.

## Introduction

Atrial fibrillation (AF) is the most common sustained cardiac arrhythmia, arising from abnormal atrial activation that produces irregular ventricular conduction and impaired hemodynamic stability [1–3]. Its pathophysiology involves atrial fibrosis, autonomic dysfunction, and abnormal calcium handling, which promote electrical and structural remodeling and increase risks of thromboembolism and multiorgan damage [4, 5]. Globally, AF affects over 37 million individuals with more than 3 million new cases annually, and its prevalence is expected to rise substantially, reaching over 12 million in the United States by 2050 and nearly 18 million in Europe by 2060 [6–8]. The economic burden is considerable, with AF-related healthcare costs exceeding $6.65 billion annually in the United States and up to €3.3 billion across Europe [9].

AF, the most common dysrhythmia in emergency departments (EDs), accounted for 143,003 visits by 113,786 patients over an 8-year period, representing 0.5% of all ED encounters [10]. AF prevalence increases sharply with age, with approximately 70% of affected individuals in the United States aged 65–85 years, and prevalence rising from 2.3% in those over 40 years to 5.9% in individuals over 65 [11]. The global prevalence of AF is expected to rise with population aging, contributing to an increasing number of patients presenting to overcrowded EDs [12, 13]. However, current management primarily targets symptom control and long-term rhythm outcomes, offering limited guidance for short-term risk stratification in acute settings, as early rhythm-control therapy reduces long-term cardiovascular events but does not substantially alter short-term hospitalization or acute risk [5, 14, 15]. In these high-volume EDs, triage remains the critical first step to identify high-risk patients requiring immediate intervention. These challenges underscore the urgent need for data-driven approaches, including machine learning-based models for predicting intensive care unit (ICU) transfer and short-term mortality, as well as real-time physiological monitoring, to enable timely risk stratification and optimize allocation of critical care resources for AF patients.

Aggarwal et al. (2023) [16] conducted a study to develop and validate a new bleeding risk prediction tool, the Direct-Acting Oral Anticoagulants (DOAC) score, for AF patients receiving direct-acting oral anticoagulants. The investigators used a multi-stage approach, including Cox proportional hazards models, to identify independent predictors. The study achieved an optimism-corrected C-statistic of 0.73, indicating a moderate predictive ability for major bleeding. Despite its ease of use, the traditional statistical approach was constrained by a limited set of preselected clinical variables and an inability to model complex, non-linear interactions, resulting in a restriction of its predictive performance in heterogeneous AF populations.

Barrett et al. (2015) [17] conducted a prospective cohort study to externally validate the RED-AF (Risk Estimator Decision Aid for Atrial Fibrillation) in ED patients with AF. Using traditional multi-variable Logistic Regression (LR), assigning points to clinical factors like age, comorbidities, symptoms, medications, and adequacy of ED rate control. It was evaluated for 30-day AF-related adverse events. In the validation cohort, the Risk Estimator Decision Aid for Atrial Fibrillation (RED-AF) achieved a c-statistic of 0.65 (95% confidence interval (CI): 0.59–0.71), also indicating only moderate discrimination and limited usefulness for high-stakes disposition decisions. These results demonstrate the performance ceiling of traditional statistical approaches for AF patients due to their inability to capture non-linear interactions and complex feature effects.

Traditionally, statistical approaches such as logistic and Cox regression have been widely used to develop clinical risk prediction models in AF. Tools such as the DOAC Score exemplify these methods, offering interpretability, transparency, and clinical familiarity, which make them suitable for explanatory analyses and routine risk stratification. However, like other regression-based systems, they typically rely on a limited number of predefined variables and assume linear relationships, which may fail to capture complex interactions underlying heterogeneous AF populations. Moreover, simplified scoring systems—while practical—often achieve only moderate discriminative performance, constraining their clinical utility in high-stakes decision making. These limitations highlight the need for more flexible, data-driven approaches that can leverage high-dimensional patient data and adapt dynamically to diverse clinical environments. In response, machine learning–based methods have gained increasing traction for their ability to model nonlinear relationships, automate feature selection, and improve predictive performance across multiple AF-related outcomes.

Machine learning methods nowadays have gained widespread application in healthcare for symptom diagnosis and patient outcome forecasting. Furthermore, they consistently demonstrate superior predictive performance compared to traditional statistical models. For instance, Rajkomar et al. (2018) [18] developed deep learning models across multiple hospitals, achieving an Area Under the Receiver Operating Characteristic curve (AUROC) of 0.85–0.86 for prolonged length of stay and outperforming traditional baselines. Similarly, Mortazavi et al. (2016) [19] reported that Random Forest (RF) delivered a 17.8% improvement in discrimination over LR for predicting 30-day readmission in heart failure patients. These successes underscore machine learning methods’ ability to capture non-linear interactions and adapt dynamically to heterogeneous patient populations. Among machine learning algorithms, tree-based ensemble methods like RF, XGBoost, LightGBM, and CatBoost have shown outstanding performance in clinical risk prediction tasks. LightGBM, in particular, offers distinct advantages: (1) High training efficiency due to histogram-based decision tree learning and optimized leaf-wise tree growing, (2) superior generalization ability while maintaining low overfitting risk, and (3) feature importance ranking with high interpretability. For example, Ke et al. (2022) [20] used LightGBM to predict early in-hospital mortality among elderly ICU patients with sepsis in MIMIC-IV and achieved AUROC = 0.870 (95% CI: 0.861-0.894). Likewise, Pan et al. (2025) [21] developed a LightGBM-based model to predict ICU mortality in patients with severe community-acquired pneumonia, achieving robust performance with an AUROC of 0.856 (95% CI: 0.792–0.921) in external validation. These results highlight LightGBM’s effectiveness in addressing key challenges such as outcome heterogeneity, non-linear feature interactions, and predictive precision—challenges that are also central to AF patients requiring critical care.

In this study, we developed a unified and systematic machine learning framework to predict ICU transfer and short-term mortality in ED patients with AF, combining rigorous data preprocessing, hybrid feature selection, multi-algorithm model development, and interpretability analyses. Our approach enhanced predictive performance, ensured generalizability, and promoted clinical relevance:

- **Model Interpretability and Clinical Integration:** We incorporated SHAP-based attribution analysis and iterative feature ablation to quantify the contribution of each predictor, providing transparent and clinically interpretable results. These strategies ensured that model predictions align with established clinical knowledge and can be meaningfully integrated into decision-making workflows.
- **Hybrid Feature Selection Strategy:**A hybrid feature selection approach was employed, combining Recursive Feature Elimination with Cross-Validation (RFECV) and Least Absolute Shrinkage and Selection Operator (LASSO). By retaining features identified by both methods, the strategy captures linear and non-linear relationships while preserving clinical relevance, resulting in a concise and informative set of 19 predictors.
- **Modular Preprocessing and Class Imbalance Mitigation:** We implemented a modular preprocessing pipeline tailored to variable types: numerical variables were median-imputed and Z-score normalized, while categorical variables were mode-imputed and one-hot encoded. To address class imbalance, the Synthetic Minority Over-sampling Technique (SMOTE) was applied, ensuring robust and stable model training across all predictive tasks.
- **Comprehensive Model Selection and Evaluation:** Six diverse machine learning algorithms—LR, RF, Gradient Boosting Machines (GBM), XGBoost, LightGBM, and CatBoost—were systematically evaluated. Model performance was assessed via stratified 5-fold cross-validation, with hyperparameters optimized through grid search, ensuring reliable, generalizable, and high-performing predictive models

## Methods

### Data Source and Study Design

This retrospective study leveraged two publicly available, de-identified electronic health record databases: MIMIC-IV (v3.1) [22] and MIMIC-IV-ED (v2.2) [23], curated by the MIT Laboratory for Computational Physiology. These datasets are derived from the Beth Israel Deaconess Medical Center (BIDMC). MIMIC-IV contains records of patients admitted to BIDMC intensive care units or the emergency department between 2008 and 2022, while MIMIC-IV-ED contains data for approximately 425,000 ED visits between 2011 and 2019. Both datasets are fully de-identified in accordance with HIPAA regulations. All data used in this study were handled in accordance with the approval of the institutional review boards, and our research team accessed the databases on 15 May 2025 after completing the required credentialing and data use agreement. Authors did not have access to any information thatune could directly identify individual participants during or after data collection.

Leveraging both MIMIC-IV-ED and MIMIC-IV datasets, we established a temporally and demographically heterogeneous ED cohort of patients with AF to improve model generalizability. Our end-to-end data science workflow systematically integrated cohort selection, exclusion of ineligible admissions, rigorous data preprocessing, extraction of clinical and laboratory features guided by domain knowledge, and model development with internal validation. The complete pipeline, detailed in Algorithm 1, ensures reproducibility and robust evaluation across diverse patient subgroups.

#### Algorithm 1 Data Preparation and Cohort Selection for Predicting Outcomes in AF Patients

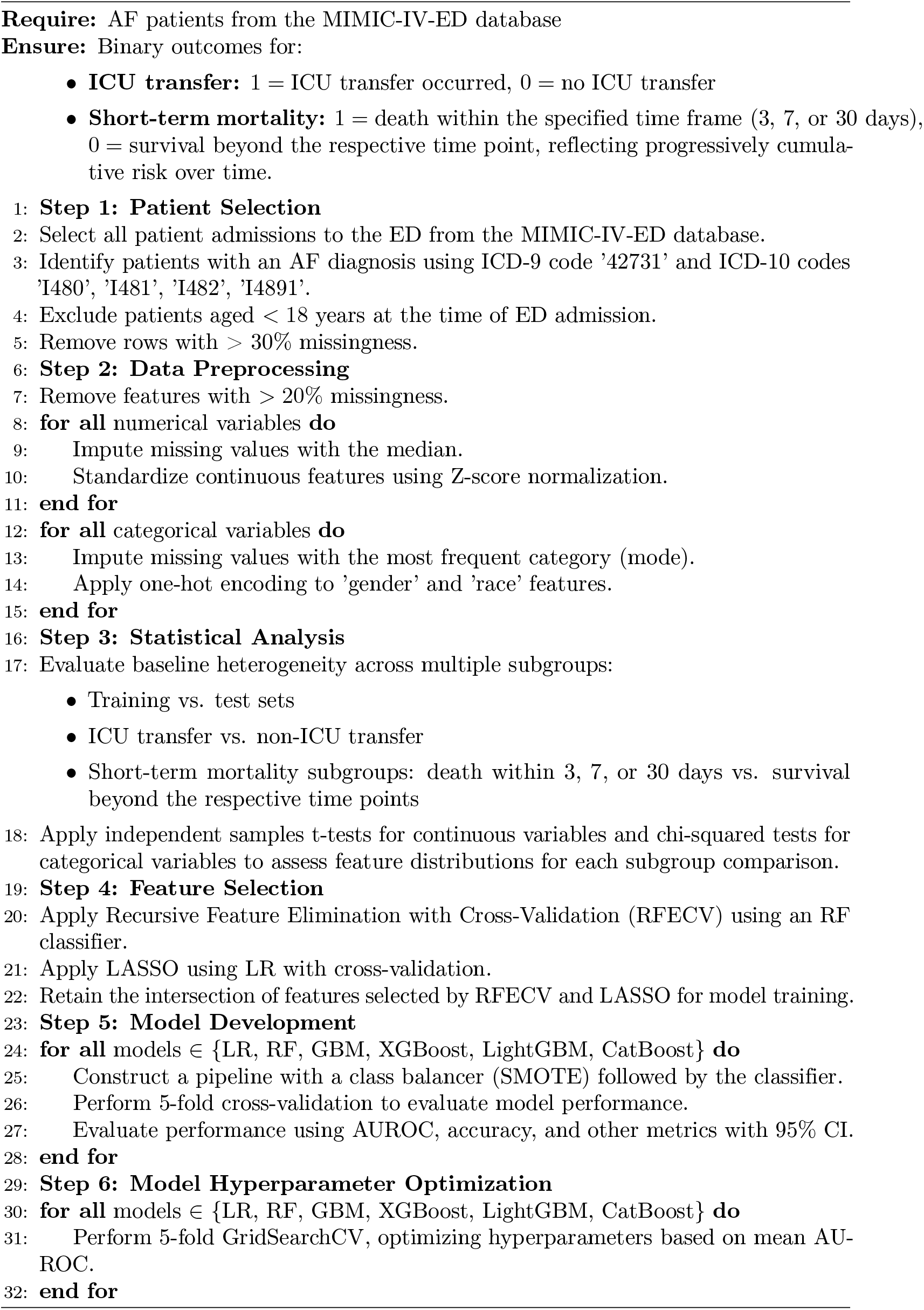
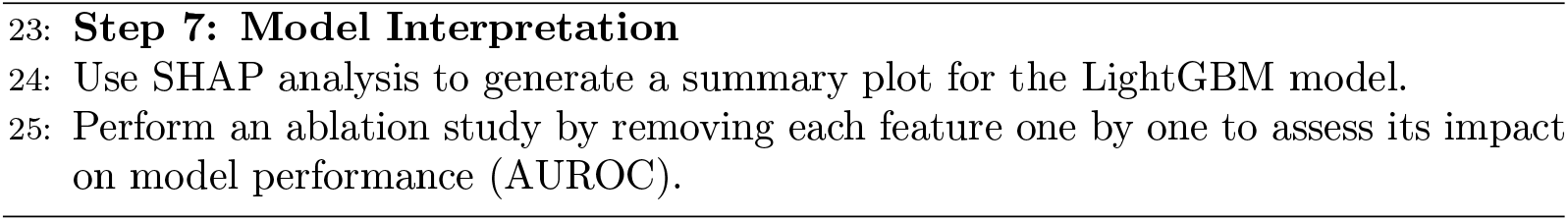

### Patient Selection

The patient cohort was established by systematically identifying all individuals with an ED admission from the MIMIC-IV-ED dataset. Data extraction for relevant patient demographics, diagnoses, vital signs, and laboratory tests was performed using structured SQL queries on the Google BigQuery platform, which allowed for efficient and reproducible data retrieval from the core MIMIC-IV database modules. The following criteria were then applied to construct the final analytical cohort: (1) Patients with a confirmed primary diagnosis of AF were included. This was identified using International Classification of Diseases, Ninth Revision (ICD-9) code ‘42731’ and International Classification of Diseases, Tenth Revision (ICD-10) codes ‘I480’, ‘I481’, ‘I482’, and ‘I4891’. (2) Patients aged less than 18 years at the time of their ED admission were excluded. The systematic application of these criteria yielded a final analytical cohort consisting of 28,951 individuals, ensuring the clinical relevance of the findings. (3) Remove rows with > 30% missingness. After applying these criteria, a final analytical cohort of 28,951 individuals was obtained. The systematic application of these criteria ensured the clinical relevance of the findings.

The predictive models were developed and evaluated using two primary categories of endpoints: ICU transfer and short-term mortality. ICU transfer was defined as a binary outcome, coded as 1 if a patient admitted through the ED subsequently had a corresponding record in the *icustays* table of the MIMIC-IV database, and 0 otherwise. Short-term mortality encompassed three temporally distinct binary outcomes—death within 3, 7, or 30 days of ED admission—capturing progressively cumulative risk over time. Specifically, 3-day mortality was defined for fatalities occurring within 72 hours of ED presentation. All mortality outcomes were systematically derived by linking patient admission and discharge information from the MIMIC-IV-ED dataset with comprehensive timestamps and clinical records from the core MIMIC-IV database, ensuring accurate temporal alignment and robust endpoint definition.

The detailed patient selection and data extraction process is depicted in Figure 1.

**Fig 1.**
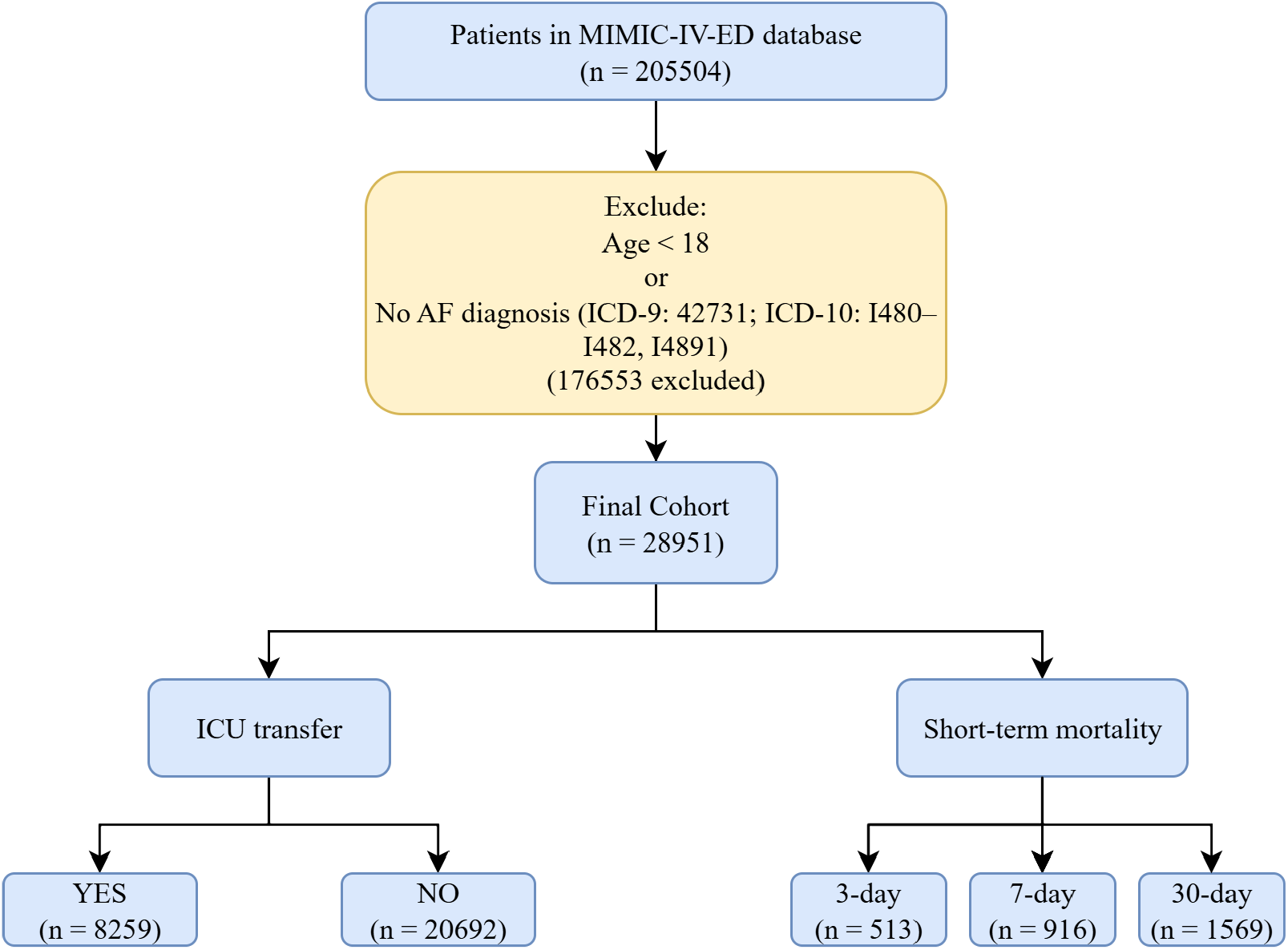
Patient Selection Process Flowchart Based on MIMIC-IV-ED and MIMIC-IV Databases.

### Data Preprocessing

A robust and systematic data preprocessing pipeline was implemented to ensure the quality, consistency, and reliability of the dataset prior to analysis. In clinical research, this step is particularly crucial given inherent data challenges such as missing values, disparate units, and varying feature scales. Proactive management of these issues is essential for mitigating bias, improving model generalizability, and enhancing interpretability, thereby ensuring that predictive models are both accurate and clinically meaningful.

To address missingness, features with more than 20% missing values were excluded from the analysis. Missing data in numerical variables were subsequently imputed using the median value [24]. This approach was selected for its simplicity and robustness, preserving the underlying data distribution without introducing complex interdependencies, which is particularly suitable for large-scale clinical datasets. Following imputation, all continuous features were standardized via z-score normalization to harmonize their measurement units and facilitate comparability. Specifically, to prevent data leakage, each feature was scaled using training-set-derived statistics (mean, *µ*, and standard deviation, *σ*), ensuring that inputs were consistently centered and scaled:

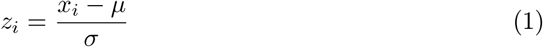

This normalization mitigates the influence of scale disparities, enhances numerical stability, and facilitates faster convergence in gradient-based optimization algorithms, ultimately improving model performance and interpretability [25].

For categorical variables, missing values were imputed using the most frequent category (mode) [24]. This strategy maintains the underlying distribution and mitigates potential imputation bias. Furthermore, the ‘gender’ and ‘race’ features were transformed using one-hot encoding, creating binary indicator variables for each category. This approach converts nominal variables into a numerical format while preserving feature matrix rank, ensuring stable coefficient estimation and interpretability.

Collectively, these type-specific preprocessing steps facilitate robust model training, enhance the performance of downstream machine learning algorithms, and provide a stable foundation for clinically meaningful and generalizable predictive modeling.

An overview of the full data preprocessing pipeline is shown in Figure 2.

**Fig 2.**
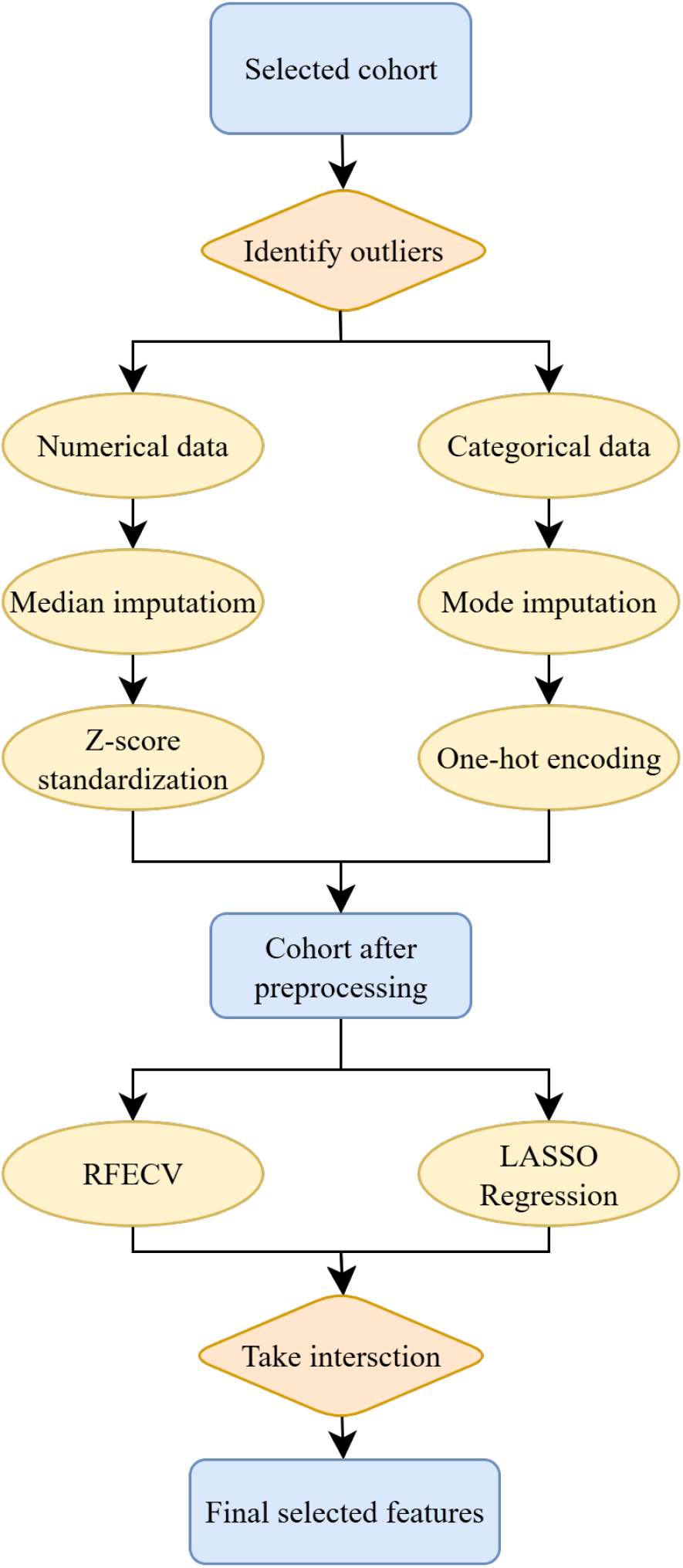
Data Preprocessing and Feature Selection Pipeline Flowchart.

### Statistical Analysis

Baseline characteristics were compared across multiple subgroups to assess potential distributional differences. Specifically, we examined feature distributions between the training and test sets, as well as across the following outcome groups: patients with and without ICU transfer, and patients with and without mortality at 3, 7, and 30 days. For continuous variables, including vital signs, were compared using two-sample Student’s t-tests under the assumption of approximate normality; this method is generally robust to modest violations of the normality assumption. [26]. For categorical variables such as comorbidities and demographic features, Pearson’s chi-square tests were used to evaluate group-wise differences in frequency distributions [27]. Test selection was guided by conventional statistical assumptions regarding variable type and sample size. A two-sided *p*-value of less than 0.05 was considered indicative of statistical significance. All tests were conducted independently for descriptive comparison purposes only and were not used to inform feature selection or model training.

### Feature Selection

To derive a clinically meaningful and statistically sound feature set, candidate predictors were selected based on prior studies in AF and critical care outcome modeling [28–30], complemented by input from clinical experts. For each continuous physiological and laboratory measure during the initial ED admission, values were calculated, as these data are crucial for the rapid assessment of patient acuity and prognosis. The final feature set comprised 80 variables grouped into three major categories:

- **Demographics and Administrative Indicators:** This group included age, gender, and race, capturing essential baseline patient characteristics known to influence disease trajectory.
- **Vital signs:** Acute physiological status was assessed using vital signs and triage data recorded at the time of ED admission, including temperature, heart rate, systolic blood pressure (SBP), diastolic blood pressure (DBP), respiratory rate (resprate), oxygen saturation (O_2_sat), pain score, and acuity.
- **Comorbidities:** Chronic conditions with established relevance to AF outcomes were incorporated. This group included congestive heart failure, peripheral vascular disease, chronic pulmonary disease, liver disease, peptic ulcer disease, rheumatic disease, hypertension, ischemic heart disease, heart failure, cerebrovascular disease, dementia, diabetes, renal disease, severe liver disease, and cancer. The Charlson Comorbidity Index was also included to provide a composite measure of comorbidity burden.

To enhance data integrity and support robust model development, variables exhibiting over 20% missingness were identified during the preprocessing stage. However, this filtering step did not result in the exclusion of any variables, thus preserving all 27 derived features for subsequent analysis.

In parallel, we performed LASSO using an LR classifier with cross-validation. The final feature subset was identified by taking the intersection of the features selected by both RFECV and LASSO. This combined approach ensured a balance between predictive power and model simplicity, resulting in a concise and transparent set of 19 features for subsequent model development.

To refine the feature set, we employed a hybrid selection strategy combining model-driven and explanation-based approaches to improve both performance and interpretability. RFECV was performed using an RF classifier, selected for its robustness to feature scaling, tolerance of multicollinearity, and ability to quantify feature importance. The RFECV procedure iteratively removed low-importance variables while optimizing five-fold cross-validated performance. Feature importance was measured by summing the total reduction in Gini impurity attributed to each feature across all nodes in the ensemble, formally defined as:

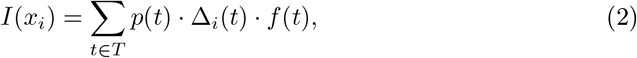

where *I*(*x*_*i*_) quantifies the contribution of feature *x*_*i*_ across the forest; *T* denotes the set of all tree nodes; *p*(*t*) is the fraction of instances routed through node *t*; Δ_*i*_(*t*) indicates the reduction in impurity attributed to *x*_*i*_ at node *t*; and *f* (*t*) reflects how often *x*_*i*_ is selected for splitting at that node.

In parallel, we performed LASSO using an LR classifier with cross-validation. This method effectively imposes a penalty that can shrink the coefficients of less informative features to zero, thereby automatically performing feature selection based on linear relationships. This approach, by identifying features with strong and direct linear associations with the outcome, serves as a robust complement to RFECV. The final feature subset was identified by taking the intersection of the top-ranked features selected by both RFECV and LASSO. This combined strategy leverages the strengths of both a non-linear, tree-based method and a linear, coefficient-based approach, resulting in a concise and transparent set of 19 predictors for subsequent model development.

The final feature subset was identified by taking the intersection of the features selected by both RFECV and LASSO. This combined approach ensured a balance between statistical significance and clinical interpretability, resulting in a concise and transparent set of 19 features for subsequent model development.

These 19 features spanned key clinical domains shown in Table 1, encompassing demographics, vital signs, and comorbidities. Specifically, the set included demographics such as age, gender, and race; vital signs including temperature, heart rate, resprate, SBP, DBP, O_2_sat, and acuity; and comorbidities such as hypertension, congestive heart failure, heart failure, renal disease, chronic pulmonary disease, rheumatic disease, and the Charlson Comorbidity Index. Collectively, these features encapsulated clinically meaningful signals critical for accurately predicting the risk of ICU transfer and mortality among AF patients.

**Table 1.**
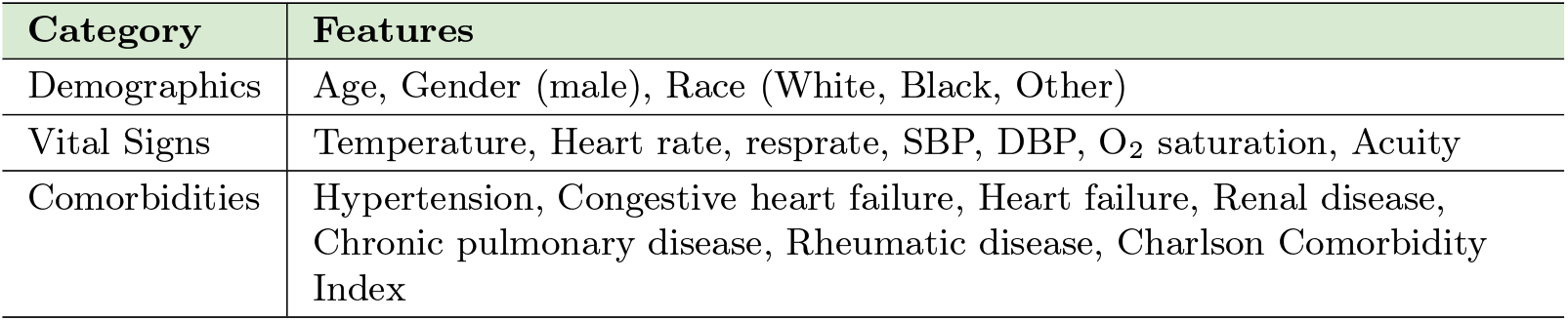
Selected Feature Set for Risk Prediction in AF Patients.

| Category | Features |
| --- | --- |
| Demographics | Age, Gender (male), Race (White, Black, Other) |
| Vital Signs | Temperature, Heart rate, resprate, SBP, DBP, O<sub>2</sub> saturation, Acuity |
| Comorbidities | Hypertension, Congestive heart failure, Heart failure, Renal disease, Chronic pulmonary disease, Rheumatic disease, Charlson Comorbidity Index |

### Model Development and Evaluation

For the purpose of predicting outcomes in AF patients after ED admissions, a supervised learning model was constructed based on the 19 selected clinical features. The dataset was evaluated using stratified 5-fold cross-validation to preserve the outcome distribution and mitigate sampling variability. This robust evaluation strategy ensures that the predictive performance is stable and reliable across different subsets of the data.

A set of six diverse machine learning algorithms, selected for their varied methodologies and practical strengths, were implemented and assessed: LR, RF, GBM, XGBoost, LightGBM, and CatBoost. For each model, a standardized pipeline was constructed comprising feature preprocessing, class imbalance correction using the Synthetic Minority Over-sampling Technique (SMOTE) oversampling algorithm [31], and model training. Model performance was assessed through stratified 5-fold cross-validation, evaluating metrics such as AUROC, accuracy, sensitivity, specificity, F1-score, positive predictive value (PPV), and negative predictive value (NPV). Confidence intervals at the 95% level were derived using bootstrap resampling. The comprehensive modeling pipeline is depicted in Figure 3.

**Fig 3.**
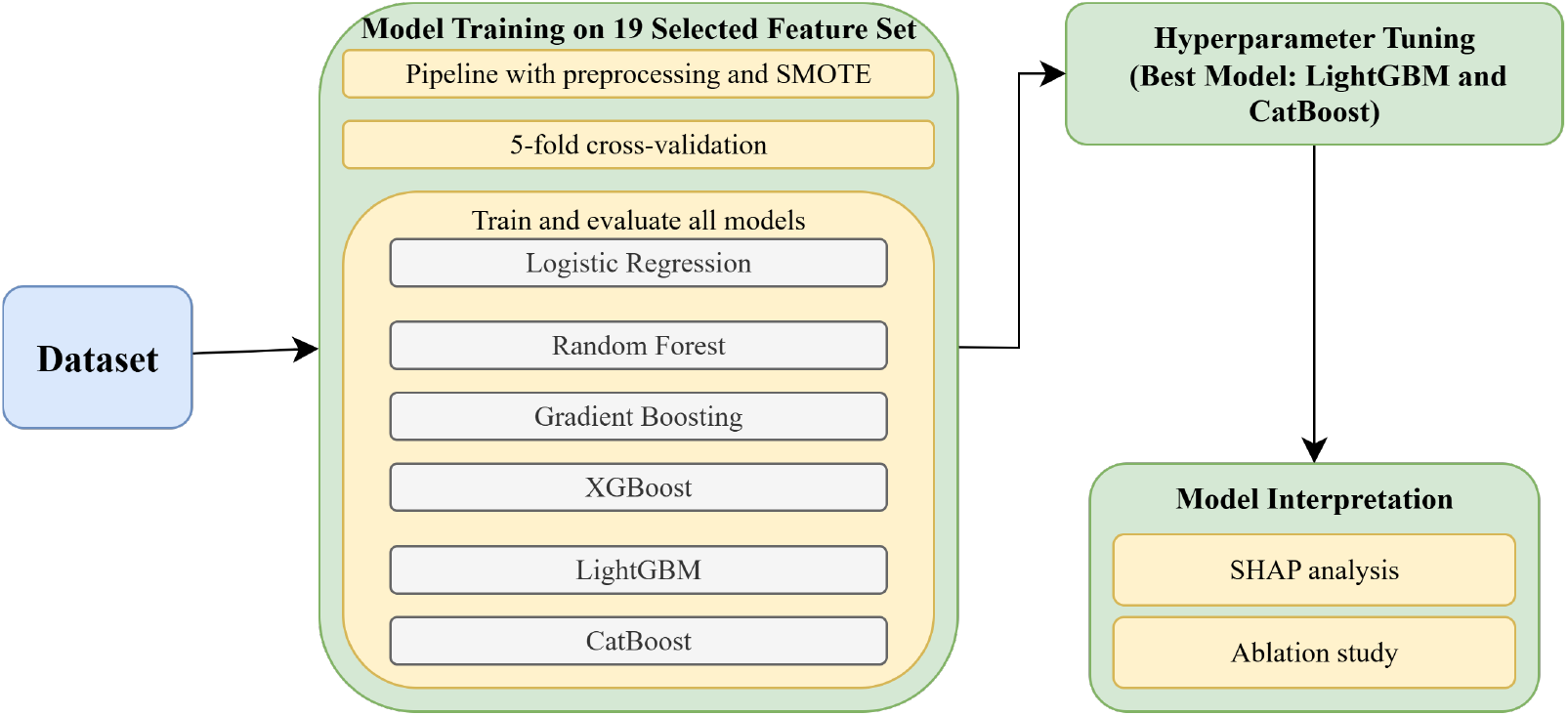
Model Training and Evaluation Workflow Flowchart Detailing Embedded Feature Selection, Final Model Optimization, and Interpretation.

Each model was chosen to reflect complementary algorithmic strengths and to capture the diverse patterns inherent in heterogeneous clinical data. LR was included as a baseline due to its simplicity, interpretability, and established use in clinical research. Its linear nature and probabilistic output make it particularly useful for risk stratification tasks. RF was implemented for its ability to capture complex non-linear relationships and interactions among features. As an ensemble method, RF aggregates the predictions of multiple decision trees, which enhances its robustness and reduces the risk of overfitting, making it a powerful tool for high-dimensional clinical data.

We also included three gradient boosting algorithms—GBM, XGBoost, and LightGBM—each known for its strong empirical performance. GBM provides a flexible boosting framework and serves as a traditional benchmark. XGBoost improves upon standard boosting with regularized loss functions and second-order gradient optimization, making it adept at capturing subtle feature interactions and controlling overfitting. LightGBM leverages histogram-based feature binning and a leaf-wise growth strategy to accelerate training while maintaining strong predictive performance, particularly in sparse or high-dimensional datasets.

CatBoost was additionally incorporated for its advanced handling of categorical variables through ordered boosting and its resistance to overfitting in small to medium-sized datasets. This characteristic made it especially well-suited for our clinical setting, which often involves variable dependencies, missing data, and nonlinear relationships.

Following the initial model evaluation, we optimized the hyperparameters for all six algorithms using a 5-fold cross-validated grid search. Key hyperparameters such as learning rate, tree depth, and regularization were tuned to optimize the mean AUROC, which was used as the primary criterion for model selection. Across all four predictive tasks, the LightGBM and CatBoost models consistently demonstrated the best performance.

Final model performance was evaluated using AUROC, with 95% CIs estimated from 2,000 bootstrap replicates to quantify statistical uncertainty and generalizability.Let *ŷ*_*i*_ be the predicted probability for sample *i* and *y*_*i*_ the true label. CatBoost was trained by minimizing the following regularized objective:

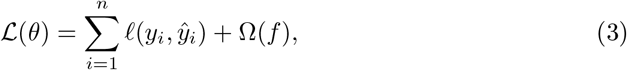

where *ℓ* is the binary cross-entropy loss function, and Ω(*f*) is a penalty term to control model complexity and prevent overfitting. This approach aligns with common gradient-boosted tree methodologies and promotes both accuracy and robustness in complex clinical datasets.

The systematic modeling framework enabled consistent comparison across classifiers, effectively addressed class imbalance and noise, and produced robust, interpretable models for stratifying ICU transfer and short-term mortality risk in ED patients with AF. By leveraging clinically meaningful features and ensuring transparency through SHAP analysis, the framework supports early, time-sensitive risk stratification and informed clinical decision-making in the emergency care setting.

### Model Interpretability and Feature Contribution Analysis

To enhance interpretability and ensure clinical plausibility, we conducted a two-pronged feature evaluation strategy comprising SHAP-based attribution analysis and iterative ablation testing.

First, we utilized SHAP [32] to evaluate the contribution of individual features to the model’s predictions. Grounded in cooperative game theory, SHAP values provide additive attributions quantifying both the direction and strength of each feature’s influence. Mathematically, the SHAP value for feature *i* is expressed as:

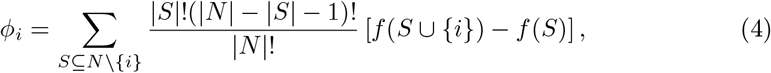

where *ϕ*_*i*_ represents the average marginal effect of adding feature *i* across all subsets *S* that exclude it. Global feature importance was visualized using SHAP summary plots, highlighting the top 19 features by mean absolute SHAP value. These findings aligned well with established clinical knowledge, emphasizing key predictors from the vital signs and comorbidity categories. For all four outcomes, features such as O_2_sat, age, acuity, and resprate consistently ranked among the most impactful predictors. The relative importance and contribution of other features, such as heart rate, blood pressure, and specific comorbidities, varied slightly across the different predictive tasks.

Second, we conducted an ablation study to evaluate the contribution and stability of each feature. Beginning with the LightGBM and CatBoost models incorporating all 19 features, we sequentially excluded one feature per iteration, retraining the model to measure the resulting change in performance. The difference in AUROC after excluding feature *i* was calculated as:

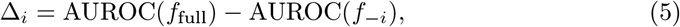

where *f*_full_ represents the model trained with the complete feature set, and *f*_−*i*_ is the model trained without feature *i*. Features whose removal led to AUROC improvement were permanently dropped, with the refined model serving as the basis for subsequent iterations. This iterative pruning proceeded until no further performance gains were achieved. The ablation outcomes were depicted through impact plots, providing clear visualization of feature importance for classification accuracy.

Together, the SHAP evaluation and systematic feature ablation enhanced the interpretability of the final model while confirming the essential role of key clinical predictors. These explainability methods bolster model transparency and reinforce its suitability for application in critical clinical decision-making settings.

## Results

### Study Cohort Profile and Comparative Statistics

The study cohort consisted of 28,951 ED patients diagnosed with AF, who were randomly divided into training (80%) and testing (20%) cohorts to support model development and validation. Among the overall population, 8,259 patients (28.5%) required transfer to the ICU, while 20,692 (71.5%) were managed without ICU escalation. Mortality outcomes occurred in 513 patients (1.8%) within 3 days, 916 patients (3.2%) within 7 days, and 1,569 patients (5.4%) within 30 days of admission. These event rates are consistent with prior literature on AF populations in critical care settings, underscoring the clinical relevance of the study endpoints. Collectively, the primary outcome measures for subsequent analyses included ICU transfer as well as all-cause mortality at 3, 7, and 30 days.

The primary objective of the comparative analysis was to assess whether systematic differences existed between the training and testing cohorts across the full set of baseline clinical features. Establishing distributional equivalence is essential to ensure that subsequent analyses are not confounded by data-split biases. No meaningful imbalances were observed across baseline characteristics (Table 2). Continuous variables—including age, SBP, DBP, heart rate, resprate, temperature, O_2_sat, and the Charlson comorbidity index—showed no statistically significant differences (*p >* 0.05 for all), confirming cohort comparability on physiologic and comorbidity profiles. For categorical variables, many exhibited low variability (e.g., sex, race, and several comorbidities) and were therefore omitted from tabular presentation to improve clarity; nevertheless, all such variables were systematically examined and confirmed to have consistent distributions between training and test cohorts. This descriptive evidence indicates that the random partitioning procedure yielded statistically balanced cohorts, supporting the validity of subsequent comparative and predictive analyses.

**Table 2.** Comparison of Feature Distributions between Training and Test Sets.

| Feature | Training Set | Test Set | P-value |
| --- | --- | --- | --- |
| resprate | 18.43 ± 3.43 | 18.35 ± 3.20 | 0.1063 |
| dbp | 75.82 ± 153.24 | 72.83 ± 22.36 | 0.1392 |
| age | 76.60 ± 12.14 | 76.35 ± 12.30 | 0.1583 |
| sbp | 132.73 ± 82.30 | 132.43 ± 24.57 | 0.7863 |
| temperature | 97.97 ± 6.43 | 97.99 ± 2.06 | 0.8049 |
| heartrate | 84.09 ± 20.62 | 84.03 ± 20.97 | 0.8437 |
| charlson_comorbidity_index | 3.56 ± 3.72 | 3.55 ± 3.74 | 0.9101 |
| O<sub>2</sub>sat | 97.48 ± 12.14 | 97.50 ± 12.19 | 0.9124 |

To further characterize the cohort, we performed comparative analyses of the full set of baseline clinical features across outcome groups, focusing first on ICU transfer. Table 3 summarizes the distributions between patients who required ICU transfer and those managed outside the ICU. Continuous variables—including age, heart rate, resprate, SBP, DBP, O_2_sat, and the Charlson comorbidity index—showed significant differences, with transfer patients generally presenting with higher heart rate, elevated resprate, greater comorbidity burden, and lower SBP, DBP, and O_2_sat. Categorical variables such as race, acuity, and comorbidities (e.g., congestive heart failure, chronic pulmonary disease, and renal disease) also demonstrated notable distributional differences, whereas others (e.g., gender, rheumatic disease, hypertension) exhibited limited variability. Some categorical variables with near-uniform distributions were labeled low variability and confirmed balanced, but are omitted for brevity. Collectively, these descriptive comparisons delineate the baseline heterogeneity between patients requiring ICU transfer and those managed without escalation, providing an epidemiological foundation for understanding outcome patterns in the cohort.

**Table 3.** Comparison of Feature Distributions between Non-ICU and ICU Transfer Groups.

| Feature | Non-ICU | ICU Transfer | P-value |  |
| --- | --- | --- | --- | --- |
| age | 76.85 ± 12.23 | 75.81 ± 12.00 | 0.0000 | |
| heartrate | 82.81 ± 20.36 | 87.24 ± 21.18 | 0.0000 | |
| resprate | 18.16 ± 2.85 | 19.05 ± 4.39 | 0.0000 | |
| sbp | 134.77 ± 86.52 | 127.41 ± 24.96 | 0.0000 | |
| charlson_comorbidity_index | 3.43 ± 3.65 | 3.89 ± 3.89 | 0.0000 | |
| O<sub>2</sub>sat | 97.64 ± 12.76 | 97.08 ± 10.46 | 0.0003 | |
| dbp | 76.88 ± 161.88 | 71.07 ± 23.00 | 0.0012 | |
| temperature | 97.97 ± 6.70 | 97.98 ± 2.55 | 0.9261 | |
| gender | – | – | 0.0007 |  |
| race | – | – | 0.0000 |  |
| congestive_heart_failure | – | – | 0.0000 |  |
| chronic_pulmonary_disease | – | – | 0.0000 |  |
| heumatic_disease | – | – | 0.4100 |  |
| hypertension | – | – | 0.4220 |  |
| heart_failure | – | – | 0.0000 |  |
| renal_disease | – | – | 0.3084 |  |
| acuity | – | – | 0.0000 |  |

Table 4 extends this comparative framework to short-term mortality outcomes within 3, 7, and 30 days. Continuous variables—including age, vital signs (heart rate, resprate, SBP, DBP, O_2_sat), and Charlson comorbidity index—exhibited statistically significant differences across outcome groups, with deceased patients generally showing older age, higher heart rate and resprate, lower SBP, and higher comorbidity burden. Categorical variables such as race, acuity, and selected comorbidities (e.g., congestive heart failure, chronic pulmonary disease, and heart failure) also demonstrated notable distributional differences, with effect magnitudes tending to increase over longer follow-up periods. As with the ICU transfer analysis, variables labeled as low variability were inspected for consistency but omitted from presentation to maintain clarity. These descriptive analyses characterize the baseline variation across mortality outcomes and provide an epidemiological context for short-term mortality.

**Table 4.** Comparison of Feature Distributions Across Short-term Mortality Outcomes (3d, 7d, 30d)

| Feature | Survived |  |  | Deceased |  |  | P-value |  |  |
| --- | --- | --- | --- | --- | --- | --- | --- | --- | --- |
|  | 3d | 7d | 30d | 3d | 7d | 30d | 3d | 7d | 30d |
| age | 76.48 ± 12.18 | 76.42 ± 12.19 | 76.38 ± 12.21 | 80.56 ± 11.05 | 80.77 ± 10.64 | 79.63 ± 10.99 | 0.0000 | 0.0000 | 0.0000 |
| heartrate | 84.05 ± 20.74 | 84.00 ± 20.76 | 83.90 ± 20.77 | 85.32 ± 17.67 | 86.35 ± 18.45 | 87.16 ± 19.05 | 0.1684 | 0.0007 | 0.0000 |
| resprate | 18.40 ± 3.37 | 18.38 ± 3.35 | 18.36 ± 3.32 | 19.16 ± 4.12 | 19.35 ± 4.26 | 19.35 ± 4.24 | 0.0000 | 0.0000 | 0.0000 |
| sbp | 132.81 ± 75.03 | 132.91 ± 75.50 | 133.15 ± 76.30 | 124.98 ± 21.73 | 125.31 ± 23.50 | 124.41 ± 23.31 | 0.0182 | 0.0024 | 0.0000 |
| charlson_comorbidity_index | 3.55 ± 3.72 | 3.54 ± 3.71 | 3.51 ± 3.69 | 3.71 ± 3.92 | 4.03 ± 4.13 | 4.38 ± 4.24 | 0.3547 | 0.0001 | 0.0000 |
| O<sub>2</sub>sat | 97.49 ± 12.25 | 97.50 ± 12.34 | 97.52 ± 12.47 | 96.88 ± 3.44 | 96.87 ± 3.25 | 96.88 ± 3.19 | 0.2615 | 0.1241 | 0.0419 |
| dbp | 75.34 ± 138.65 | 75.43 ± 139.63 | 75.60 ± 141.27 | 68.97 ± 12.99 | 68.93 ± 13.58 | 68.73 ± 14.07 | 0.2986 | 0.1592 | 0.0542 |
| temperature | 97.98 ± 5.86 | 97.98 ± 5.90 | 97.98 ± 5.95 | 97.60 ± 3.92 | 97.78 ± 3.00 | 97.84 ± 2.83 | 0.1433 | 0.3060 | 0.3309 |
| gender | — | — | — | — | — | — | 0.4565 | 0.0997 | 0.6761 |
| race | — | — | — | — | — | — | 0.0000 | 0.0000 | 0.0000 |
| congestive_heart_failure | — | — | — | — | — | — | 0.1102 | 0.9671 | 0.0072 |
| chronic_pulmonary_disease | — | — | — | — | — | — | 0.8232 | 0.2461 | 0.0024 |
| rheumatic_disease | — | — | — | — | — | — | 0.2227 | 0.0567 | 0.5608 |
| hypertension | — | — | — | — | — | — | 0.6623 | 0.2048 | 0.0276 |
| heart_failure | — | — | — | — | — | — | 0.1102 | 0.9671 | 0.0072 |
| renal_disease | — | — | — | — | — | — | 0.1234 | 0.5369 | 0.0233 |
| acuity | — | — | — | — | — | — | 0.0000 | 0.0000 | 0.0000 |

Following the comparative analyses across ICU transfer and mortality outcomes, several consistent patterns emerged regarding baseline patient characteristics. Patients requiring ICU transfer or experiencing mortality within 3, 7, or 30 days were generally older and exhibited higher heart rate and resprate, lower SBP, and greater comorbidity burden as measured by the Charlson index. Categorical features such as race and selected comorbidities (e.g., congestive heart failure, chronic pulmonary disease, heart failure, and acuity) also showed distributional differences across outcome groups, although the strength and statistical significance of these associations varied over time. Collectively, these descriptive findings delineate clinically meaningful variation within the study cohort and provide essential context for subsequent predictive modeling, which formally quantifies the contribution of baseline features to patient outcomes.

### Evaluation and Comparison of Model Performance

To comprehensively evaluate the predictive performance and robustness of six machine learning classifiers across four clinically meaningful outcomes, we conducted a comparative evaluation using AUROC, accuracy, F1 score, sensitivity, specificity, PPV, and NPV, each accompanied by 95% CIs. The outcomes were grouped into two clinically relevant categories: (i) ICU transfer from the ED, and (ii) short-term mortality at 3, 7, and 30 days. Model performance was estimated through stratified 5-fold cross-validation on the training cohort and subsequently validated on an independent test cohort. The primary results are reported in Tables 5–8, which summarize model performance for ICU transfer prediction and short-term mortality prediction across 3, 7, and 30 days, while corresponding Receiver Operating Characteristic (ROC) curves are presented in Figure 4 (panels a–h).

**Table 5.** Training Set Performance of Classifiers for ICU Transfer (with 95% CI for AUROC)

| Model | AUROC (95% CI) | Accuracy | F1 Score | Sensitivity | Specificity | PPV | NPV |
| --- | --- | --- | --- | --- | --- | --- | --- |
| LR | 0.7590 (0.7555–0.7622) | 0.6610 | 0.5147 | 0.6299 | 0.6735 | 0.4351 | 0.8201 |
| RF | 0.9983 (0.9981–0.9985) | 0.9935 | 0.9888 | 0.9968 | 0.9922 | 0.9809 | 0.9987 |
| GBM | 0.7923 (0.7892–0.7956) | 0.7358 | 0.5366 | 0.5362 | 0.8155 | 0.5370 | 0.8150 |
| XGBoost | 0.7924 (0.7891–0.7958) | 0.7303 | 0.5377 | 0.5500 | 0.8022 | 0.5260 | 0.8171 |
| LightGBM | 0.8244 (0.8214–0.8274) | 0.7762 | 0.5268 | 0.4366 | 0.9118 | 0.6639 | 0.8022 |
| CatBoost | 0.7870 (0.7838–0.7903) | 0.7301 | 0.5327 | 0.5391 | 0.8064 | 0.5264 | 0.8142 |

**Fig 4.**
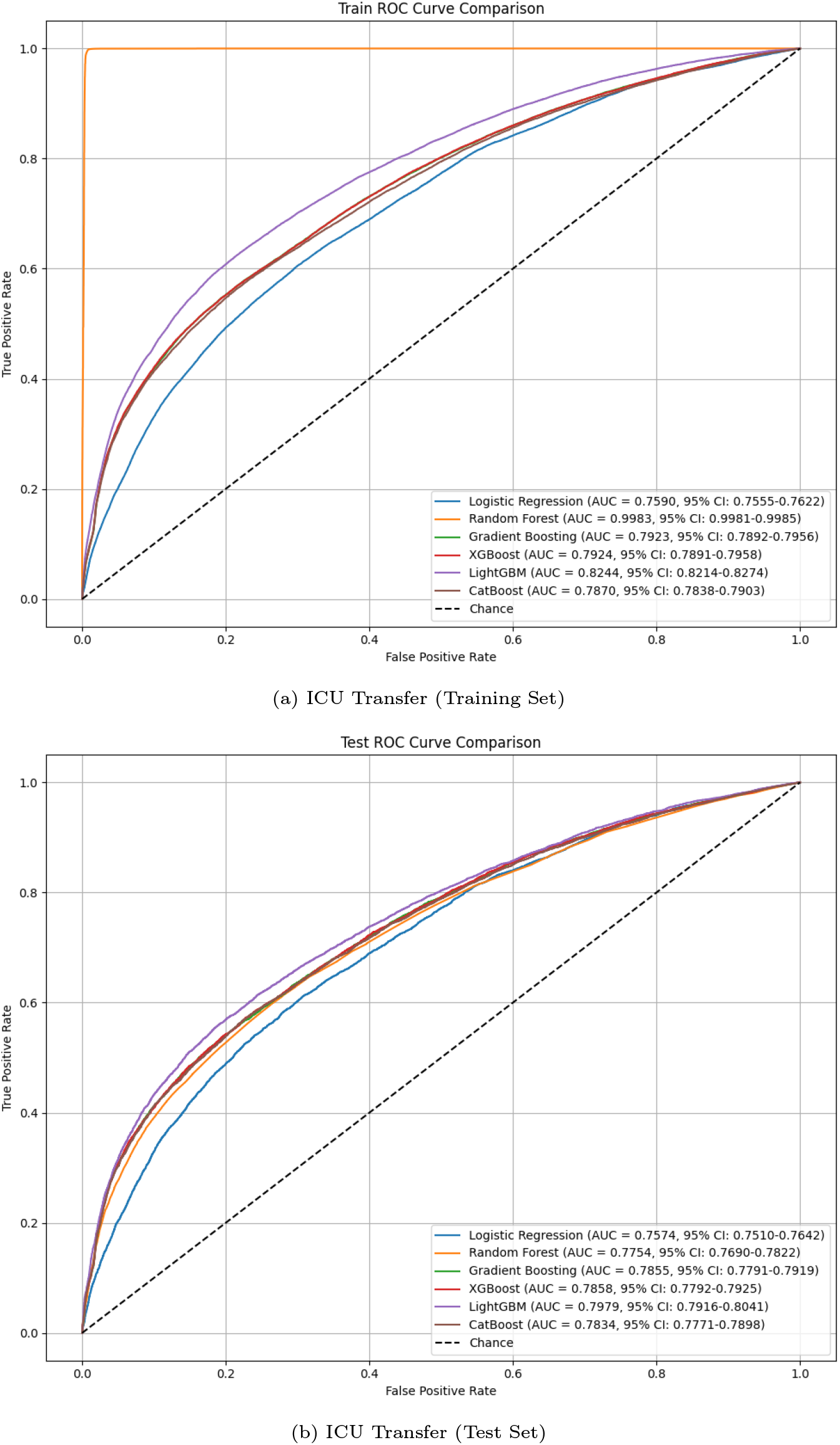
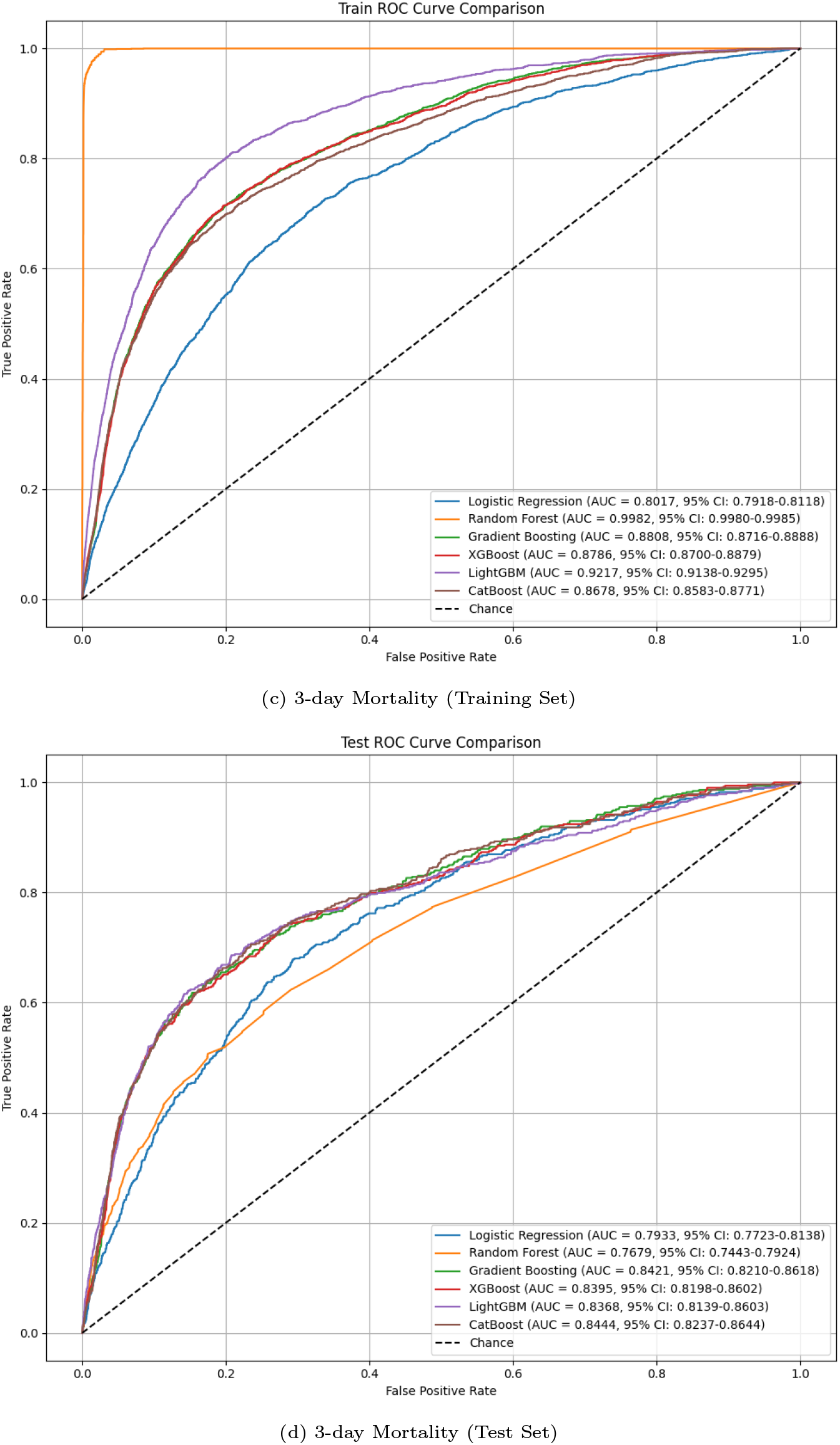
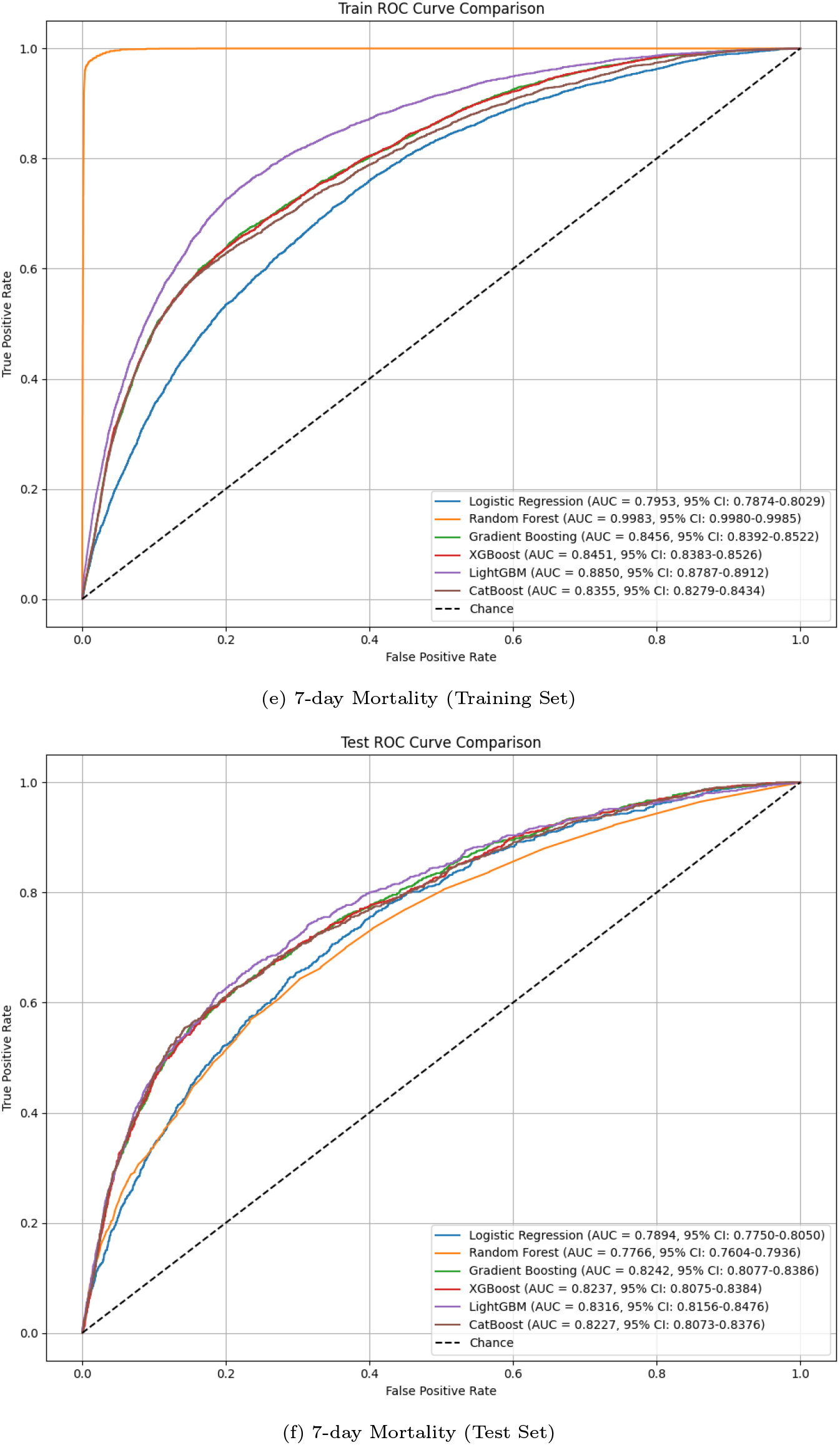
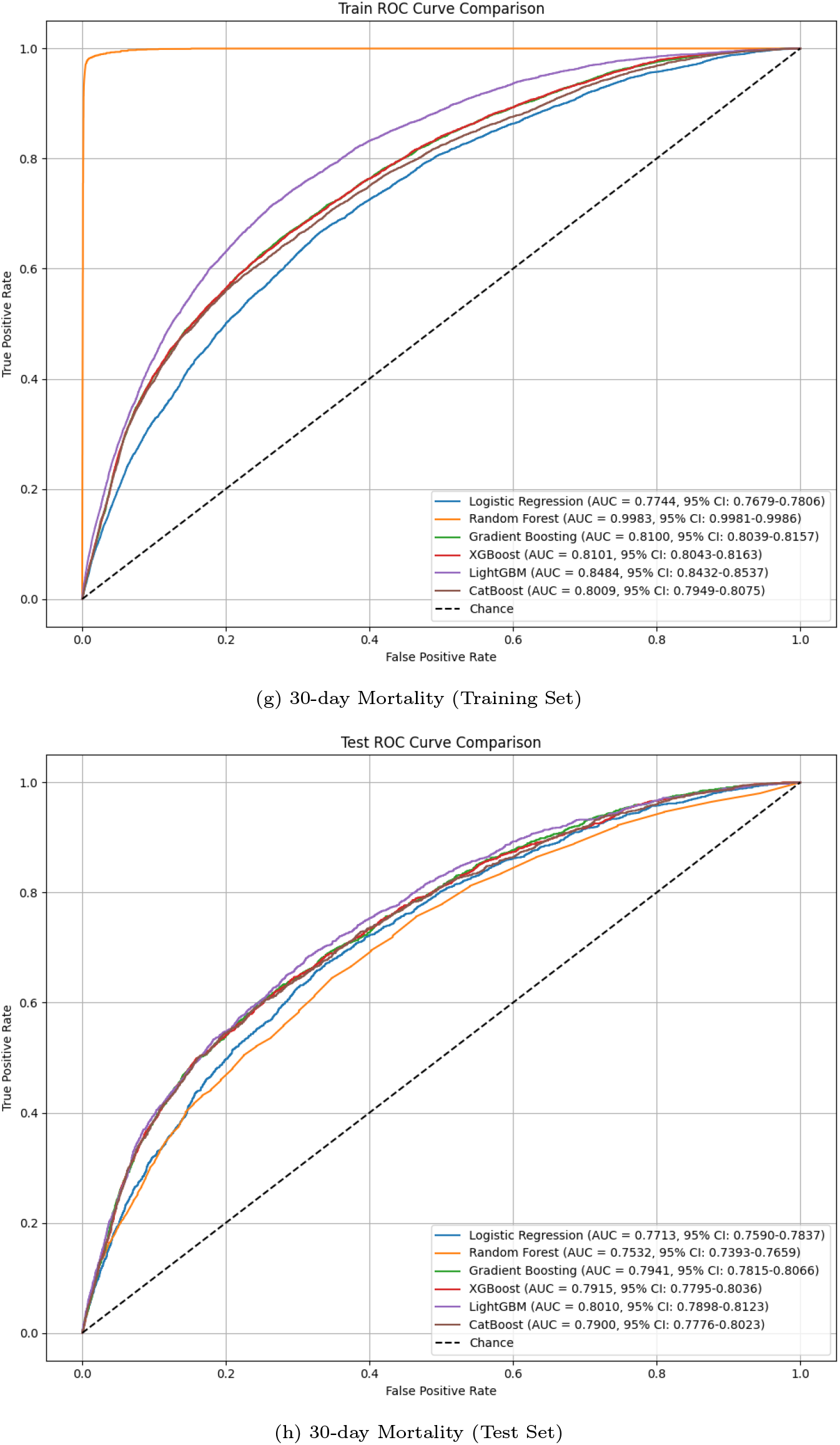
AUROC curves for training and test cohorts. Panels (a–b) show ICU transfer prediction; (c–d) 3-day mortality; (e–f) 7-day mortality; (g–h) 30-day mortality.

For ICU transfer prediction, tree-based ensemble models consistently outperformed the linear baseline. On the test set, LightGBM achieved the highest discriminative ability (AUROC: 0.7979, 95% CI: 0.7916–0.8041) with strong specificity (0.9053) and PPV (0.6399), though at the expense of lower sensitivity (0.4214), indicating a conservative profile favoring precision over recall. RF exhibited comparatively high specificity (0.8825) but very low sensitivity (0.4202), and its near-perfect performance on the training set (AUROC ≈ 0.9983) contrasted with marked declines on the test set, suggesting substantial overfitting. LR yielded the lowest performance (AUROC: 0.7574), underscoring its limitations in modeling nonlinear relationships within heterogeneous clinical data. Gradient boosting variants, including XGBoost (AUROC: 0.7858), GBM (0.7855), and CatBoost (0.7834), achieved intermediate discrimination with balanced sensitivity and specificity, though not surpassing LightGBM.

For short-term mortality prediction, LightGBM achieved the highest performance at the 7- and 30-day horizons, with AUROCs of 0.8316 and 0.8010, respectively, and consistently superior F1 scores across all time points. CatBoost attained the best AUROC for 3-day mortality (0.8444 vs. 0.8368 for LightGBM), highlighting its sensitivity to very early risk, although LightGBM maintained higher F1 scores and specificity at this horizon. Particularly at 30 days, LightGBM achieved the strongest balance of precision and recall, with the highest F1 score (0.2428) and PPV (0.2063), reflecting robustness in an imbalanced outcome setting. Gradient boosting algorithms overall (LightGBM, CatBoost, XGBoost, GBM) yielded stable discrimination (AUROC ≈ 0.79–0.84), while the RF model exhibited substantially inflated training performance but poor generalization, indicating marked overfitting. LR consistently underperformed across all horizons, underscoring the limitations of traditional statistical approaches in high-dimensional, heterogeneous clinical data.

Overall, gradient boosting models—including LightGBM, CatBoost, XGBoost, and GBM—demonstrated robust and generalizable performance across ICU transfer and short-term mortality prediction tasks. LightGBM provided consistently stable discrimination across multiple outcomes and time horizons, particularly for ICU transfer and 7- to 30-day mortality, whereas CatBoost achieved superior AUROC for 3-day mortality, reflecting heightened sensitivity to early physiological deterioration. XGBoost and GBM offered competitive discrimination with balanced sensitivity and specificity, whereas RF showed strong training performance but substantial overfitting, and LR consistently underperformed. These patterns support a layered risk stratification logic, whereby patients can be assigned to risk strata that reflect both immediate and longer-term vulnerability. Collectively, the results highlight the complementary strengths of gradient boosting models and suggest that model selection may be tailored according to the clinical time horizon of interest, with LightGBM providing the most consistent overall performance.

**Table 6.** Test Set Performance of Classifiers for ICU Transfer (with 95% CI for AUROC)

| Model | AUROC (95% CI) | Accuracy | F1 Score | Sensitivity | Specificity | PPV | NPV |
| --- | --- | --- | --- | --- | --- | --- | --- |
| LR | 0.7574 (0.7510–0.7642) | 0.6596 | 0.5129 | 0.6277 | 0.6724 | 0.4335 | 0.8189 |
| RF | 0.7754 (0.7690–0.7822) | 0.7507 | 0.4901 | 0.4202 | 0.8825 | 0.5879 | 0.7924 |
| GBM | 0.7855 (0.7791–0.7919) | 0.7305 | 0.5284 | 0.5297 | 0.8103 | 0.5271 | 0.8119 |
| XGBoost | 0.7858 (0.7792–0.7925) | 0.7261 | 0.5310 | 0.5434 | 0.7991 | 0.5193 | 0.8142 |
| LightGBM | 0.7979 (0.7916–0.8041) | 0.7674 | 0.5082 | 0.4214 | 0.9053 | 0.6399 | 0.7968 |
| CatBoost | 0.7834 (0.7771–0.7898) | 0.7269 | 0.5271 | 0.5334 | 0.8044 | 0.5211 | 0.8120 |

**Table 7.**
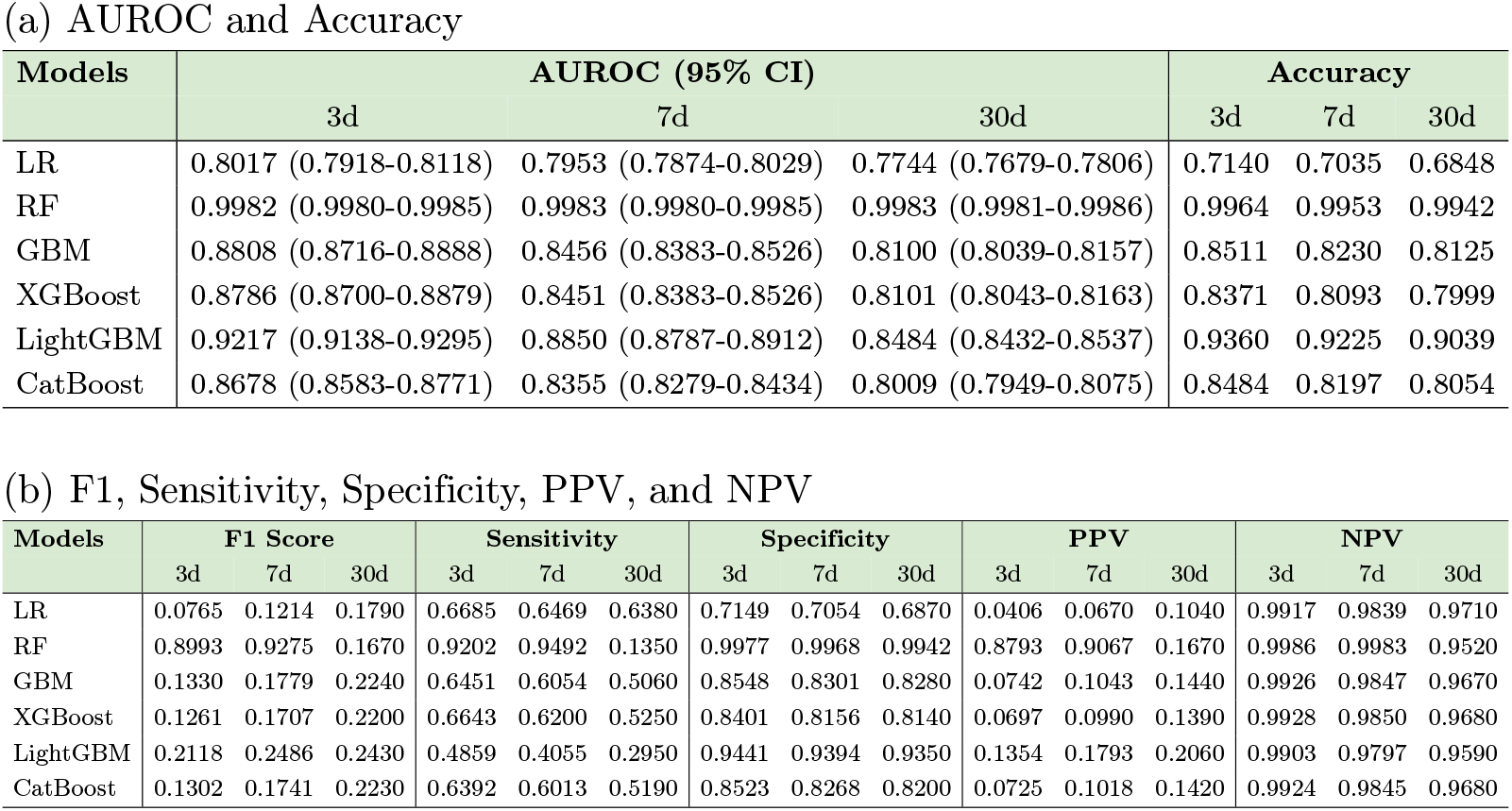
Performance comparison of machine learning models for short-term mortality prediction at 3, 7, and 30 days in the training set. (a) AUROC and Accuracy with 95% CI; (b) F1, Sensitivity, Specificity, PPV, and NPV.

**Table 8.** Performance comparison of machine learning models for short-term mortality prediction at 3, 7, and 30 days in the test set. (a) AUROC and Accuracy with 95% CI; (b) F1, Sensitivity, Specificity, PPV, and NPV.

| Models | AUROC (95% CI) |  |  | Accuracy |  |  |
| --- | --- | --- | --- | --- | --- | --- |
|  | 3d | 7d | 30d | 3d | 7d | 30d |
| LR | 0.7933 (0.7723-0.8138) | 0.7894 (0.7750-0.8050) | 0.7713 (0.7590-0.7837) | 0.7130 | 0.7030 | 0.6840 |
| RF | 0.7679 (0.7443-0.7924) | 0.7766 (0.7604-0.7936) | 0.7532 (0.7393-0.7659) | 0.9720 | 0.9550 | 0.9270 |
| GBM | 0.8421 (0.8210-0.8618) | 0.8242 (0.8077-0.8386) | 0.7941 (0.7815-0.8066) | 0.8470 | 0.8210 | 0.8110 |
| XGBoost | 0.8395 (0.8198-0.8602) | 0.8237 (0.8075-0.8384) | 0.7915 (0.7795-0.8036) | 0.8350 | 0.8070 | 0.7980 |
| LightGBM | 0.8368 (0.8139-0.8603) | 0.8316 (0.8156-0.8476) | 0.8010 (0.7898-0.8123) | 0.9320 | 0.9200 | 0.9000 |
| CatBoost | 0.8444 (0.8237-0.8644) | 0.8227 (0.8073-0.8376) | 0.7900 (0.7776-0.8023) | 0.8460 | 0.8180 | 0.8040 |

(b) F1, Sensitivity, Specificity, PPV, and NPV
| Models | F1 Score |  |  | Sensitivity |  |  | Specificity |  |  | PPV |  |  | NPV |  |  |
| --- | --- | --- | --- | --- | --- | --- | --- | --- | --- | --- | --- | --- | --- | --- | --- |
|  | 3d | 7d | 30d | 3d | 7d | 30d | 3d | 7d | 30d | 3d | 7d | 30d | 3d | 7d | 30d |
| LR | 0.0754 | 0.1212 | 0.1793 | 0.6589 | 0.6478 | 0.6379 | 0.7145 | 0.7046 | 0.6866 | 0.0400 | 0.0669 | 0.1043 | 0.9915 | 0.9839 | 0.9707 |
| RF | 0.1233 | 0.1507 | 0.1672 | 0.1098 | 0.1272 | 0.1354 | 0.9879 | 0.9817 | 0.9722 | 0.1408 | 0.1851 | 0.2185 | 0.9840 | 0.9718 | 0.9515 |
| GBM | 0.1235 | 0.1680 | 0.2242 | 0.6066 | 0.5717 | 0.5055 | 0.8517 | 0.8293 | 0.8281 | 0.0688 | 0.0985 | 0.1441 | 0.9917 | 0.9834 | 0.9669 |
| XGBoost | 0.1173 | 0.1629 | 0.2196 | 0.6202 | 0.5925 | 0.5246 | 0.8390 | 0.8144 | 0.8136 | 0.0648 | 0.0944 | 0.1389 | 0.9919 | 0.9839 | 0.9676 |
| LightGBM | 0.1717 | 0.2206 | 0.2428 | 0.3981 | 0.3582 | 0.2951 | 0.9415 | 0.9383 | 0.9350 | 0.1095 | 0.1594 | 0.2063 | 0.9886 | 0.9781 | 0.9586 |
| CatBoost | 0.1217 | 0.1686 | 0.2225 | 0.6050 | 0.5848 | 0.5186 | 0.8499 | 0.8255 | 0.8200 | 0.0677 | 0.0985 | 0.1416 | 0.9917 | 0.9839 | 0.9675 |

### Model Explanation and Clinical Insight via SHAP

To enhance clinical interpretability and assess the robustness of the LightGBM- and CatBoost-based frameworks in predicting ICU transfer and short-term mortality among patients with AF, we employed SHAP values to quantify the relative contribution of each predictor (Figure 5; panel a shows ICU transfer, panels b–d show 3-, 7-, and 30-day mortality). The analysis encompassed 19 clinically meaningful features, with global SHAP summary plots ranking them according to their mean absolute SHAP values. In these plots, the horizontal axis captures both the magnitude and direction of each feature’s influence on the model output, while the color gradient encodes the original feature values (red denoting higher and blue denoting lower values).

**Fig 5.**
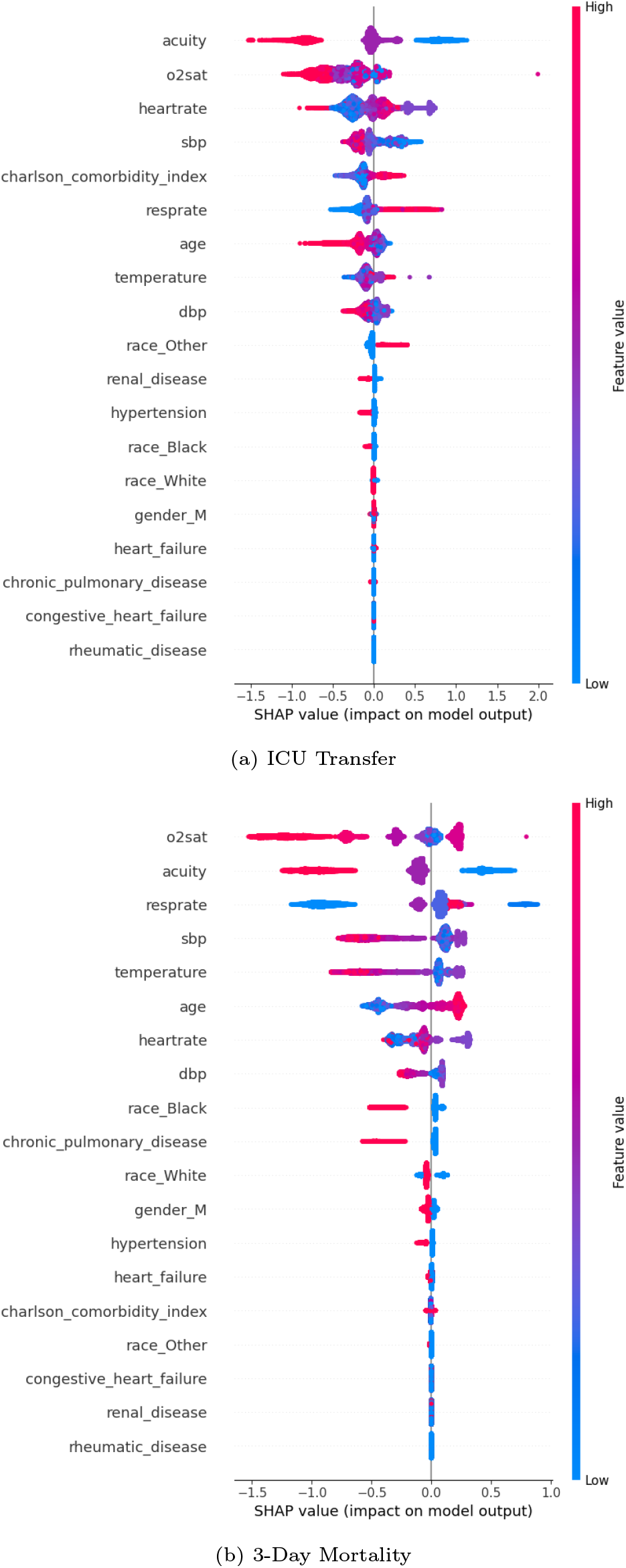
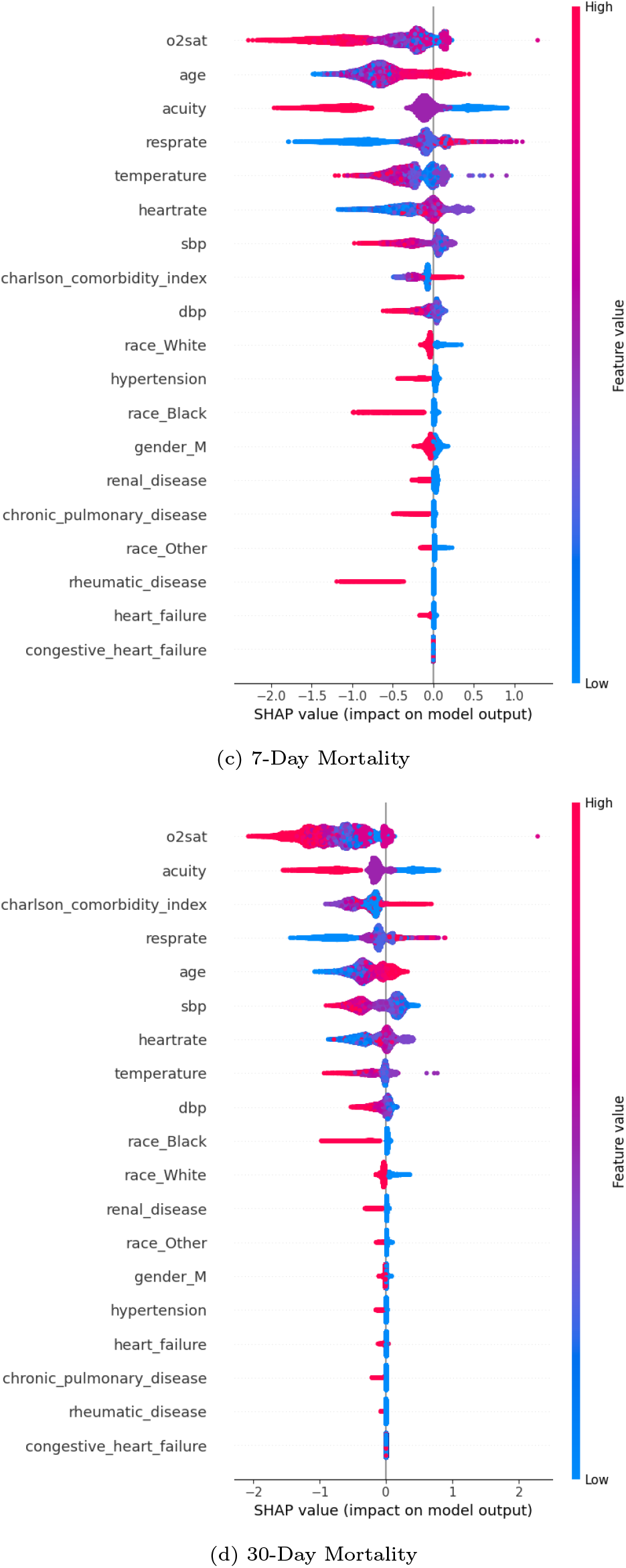
SHAP summary plots showing feature contributions and interpretability. Panel (a) shows ICU transfer; panels (b–d) show 3-, 7-, and 30-day mortality.

For ICU transfer, acuity emerged as the strongest predictor (Figure 5, panel a), surpassing all other features in relative importance. The SHAP summary plot further identified O_2_sat, heart rate, and SBP as subsequent contributors, highlighting the pivotal role of hemodynamic and respiratory stability in determining ICU transfer.

Resprate also ranked prominently, reinforcing systemic stress as a key physiological trigger for escalation of care. Importantly, surrogate post-outcome variables (e.g., length of stay) were deliberately excluded to avoid information leakage. Despite this restriction, the model maintained robust discriminative performance, underscoring the clinical validity and reliability of the identified predictors.

For short-term mortality prediction (3-, 7-, and 30-day), the SHAP summary plots (Figure 5, panels b–d) revealed O_2_sat as the dominant determinant: lower O_2_sat values consistently drove higher mortality risk, and its absence would be expected to cause the largest decline in predictive power. The contributions of age and acuity were also critical, capturing systemic frailty and acute severity; without these features, the model’s ability to detect high-risk patients would be markedly reduced. For 30-day mortality specifically, the Charlson Comorbidity Index played a notable role, highlighting the weight of chronic disease burden in shaping longer-term outcomes. Additional contributors—including SBP, resprate, and temperature—added secondary yet consistent predictive value, reinforcing their complementary role in mortality risk stratification.

Overall, the SHAP-based analysis delineates a clear and clinically coherent hierarchy of risk determinants for ICU transfer and short-term mortality in AF patients. For ICU transfer and very early (3-day) mortality, acute physiological parameters—most notably O_2_sat, respiratory rate, heart rate, and systolic blood pressure—emerged as dominant contributors, underscoring the pivotal role of hemodynamic and respiratory instability as immediate triggers of deterioration. As the prediction horizon extended to 7 and 30 days, age and the Charlson Comorbidity Index gained prominence, reflecting the cumulative effects of systemic frailty and chronic disease burden on longer-term vulnerability. This graded, additive progression highlights a stratified risk architecture in which acute physiological derangements precipitate early adverse events, while baseline vulnerability factors amplify the likelihood of later mortality. Clinically, these findings reinforce the plausibility of the model outputs and provide actionable insights: patients identified at high acute risk may benefit from timely escalation of monitoring and intervention, whereas those flagged by frailty-related features warrant sustained surveillance and tailored management strategies.

### Ablation Analysis of Model Predictive Factors

To further assess the stability and interpretability of the LightGBM- and CatBoost-based frameworks in predicting ICU transfer and short-term mortality among AF patients, we conducted a comprehensive feature ablation analysis. As shown in Figure 6, panels (a–d), each of the top 19 SHAP-ranked features was sequentially removed, and the model was retrained on the remaining feature set. Performance was evaluated using stratified five-fold cross-validation repeated ten times to ensure statistical robustness. The baseline model achieved an AUROC of 0.8444 for 3-day mortality, 0.8316 for 7-day mortality, 0.8010 for 30-day mortality, and 0.7979 for ICU transfer, which served as reference benchmarks for quantifying the marginal contribution of each feature.

**Fig 6.**
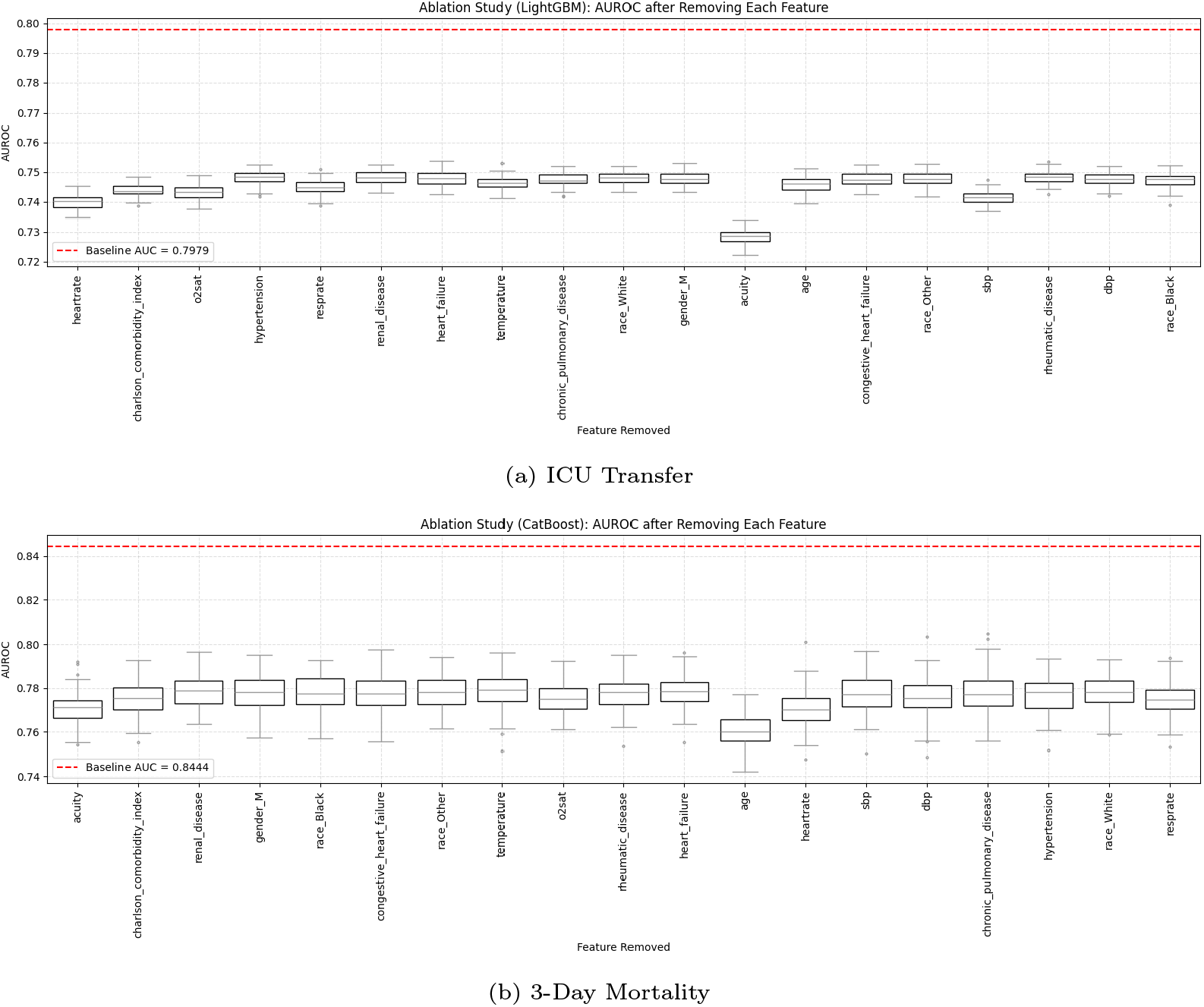
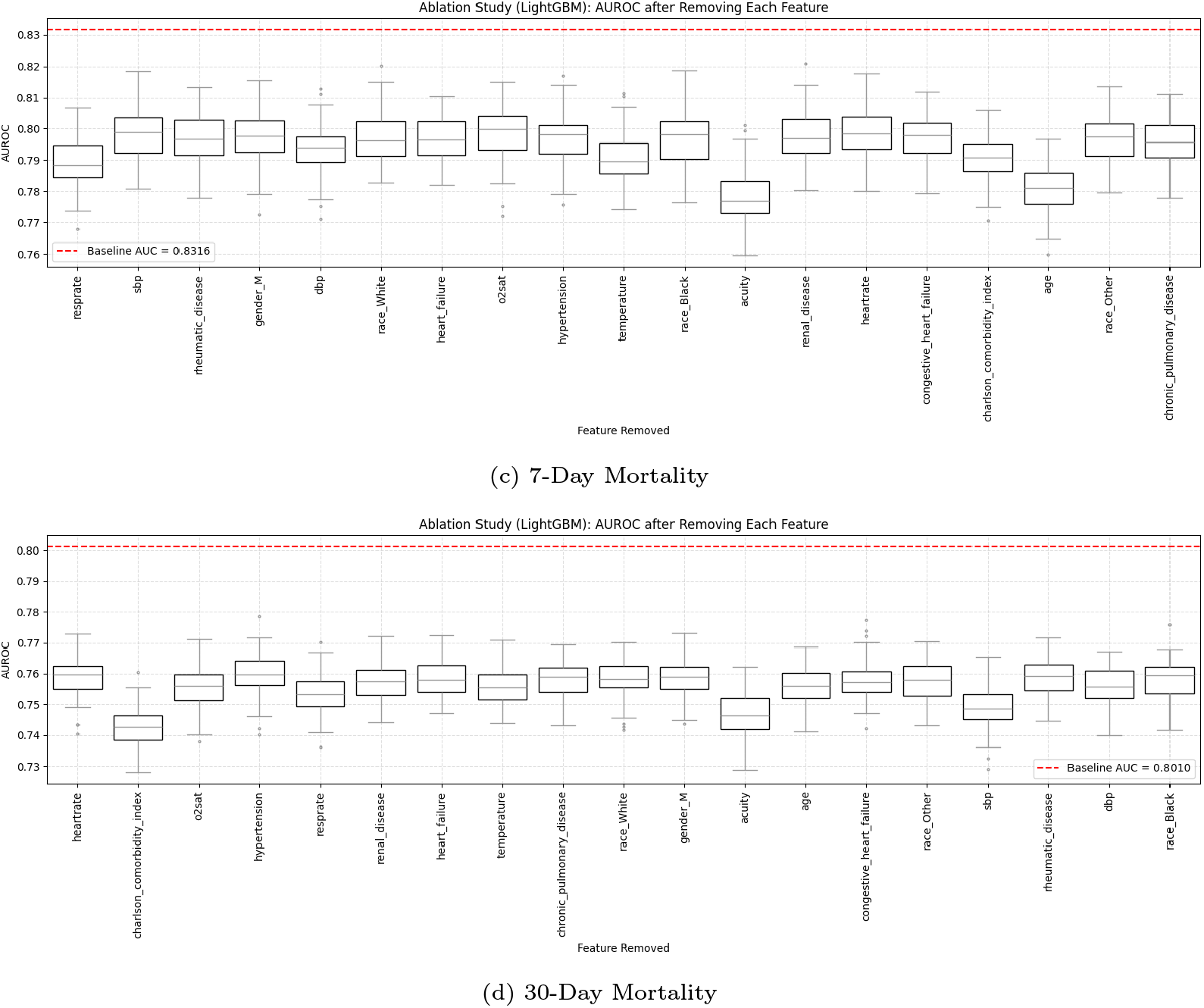
Ablation study showing the impact of removing individual features on AUROC. Panel (a) shows ICU transfer; panels (b–d) show 3-, 7-, and 30-day mortality.

For short-term mortality prediction, the exclusion of age and acuity produced the most substantial declines in AUROC, underscoring their central roles in capturing systemic deterioration and patient frailty. For 30-day mortality, a decline in AUROC due to the Charlson comorbidity index being removed is also remarkable. Similarly, for ICU transfer, the removal of acuity, heart rate, and SBP led to the largest drops, reflecting the critical importance of hemodynamic stability and acute severity in escalation-of-care decisions. Features such as temperature and resprate contributed moderately but consistently across tasks.

Taken together with the SHAP results, the ablation study shows that model performance depends on a distributed set of physiologic, demographic, and laboratory indicators, rather than on a single dominant feature. This distributed reliance enhances model resilience to missing or noisy inputs and supports its generalizability across diverse AF subgroups encountered in the emergency setting.

Clinically, the prominence of features such as O_2_sat, acuity, and SBP aligns with established risk factors for both acute deterioration and mortality in AF patients, providing physiological credibility to the LightGBM predictions. By leveraging interpretable and clinically relevant features, the model demonstrates strong potential for integration into real-time risk stratification workflows, enabling proactive identification of high-risk patients and more efficient allocation of ICU resources.

In summary, the ablation analysis confirms the stability and robustness of the LightGBM-based predictive framework. Its reliance on multiple complementary predictors enhances interpretability, statistical validity, and practical clinical utility, supporting its role in improving AF patient management in emergency and critical care settings.

## Discussion

### Summary and Review of Existing Models

This study developed robust and interpretable machine learning models to predict ICU transfer and short-term mortality in ED patients with AF, using data from the MIMIC-IV-ED and MIMIC-IV databases. A structured pipeline including missing data imputation, hybrid feature selection with RFECV and LASSO, and multi-model benchmarking was applied, resulting in a final set of 19 clinically meaningful features encompassing baseline demographics, comorbidities, and initial vital signs. LightGBM demonstrated the highest performance for ICU transfer (AUROC = 0.7979, 95% CI: 0.7916–0.8041) and for 7- and 30-day mortality (AUROC = 0.8316, 95% CI: 0.8156–0.8476; AUROC = 0.8010, 95% CI: 0.7898–0.8123), while CatBoost achieved the best performance for 3-day mortality (AUROC = 0.8444, 95% CI: 0.8237–0.8644), surpassing other gradient boosting methods. This division of strengths suggests that LightGBM provides stable and generalizable predictions across broader outcome horizons, whereas CatBoost is particularly sensitive to very early (3-day) physiological deterioration, offering complementary utility for time-specific risk stratification. SHAP-based analyses identified O_2_sat and acuity as the most influential predictors across outcomes, with resprate also playing a key role in mortality prediction, underscoring their central importance in patient risk stratification. The consistent high performance across multiple evaluation metrics and stratified cross-validation highlights the models’ discriminative power and potential generalizability for early identification of high-risk AF patients in the ED.

The findings of this study have important clinical and operational implications. First, by leveraging vital signs and other clinical features collected at the time of ED triage—captured in the MIMIC-IV-ED triage table during patients’ initial assessment upon arrival—the models enable early identification of individuals at elevated risk for ICU transfer or short-term mortality. This provides a crucial window for initiating timely interventions, such as closer monitoring, early hemodynamic support, or expedited specialist consultation. Second, the interpretability afforded by SHAP enhances clinician confidence and supports actionable decision-making at the bedside. For example, patients presenting with low O_2_sat, high acuity scores, or elevated resprate are flagged by the models as high-risk, offering transparent guidance that can inform immediate clinical responses. Third, at the healthcare system level, integrating these predictive models could aid in triaging ED resources and prioritizing care for high-risk AF patients, improving operational efficiency without necessitating additional tests or infrastructure. It is important to note that the predictions do not differentiate between deaths arising from medical complications versus those following comfort-focused care, which represent distinct clinical pathways requiring tailored interventions. Overall, the reliance on routinely collected triage and EHR(Electronic Health Record) data supports broad applicability and facilitates deployment across diverse ED settings.

This study leverages the MIMIC-IV-ED and MIMIC-IV databases to develop robust and interpretable machine learning models for predicting ICU transfer and short-term mortality in ED patients with AF. Using a hybrid feature selection approach combining RFECV and LASSO, the models achieved strong discriminative performance, with SHAP analysis confirming the relevance of key triage and clinical variables. Stratified 5-fold cross-validation and ablation studies validated model stability, highlighting their potential for early, actionable risk stratification and practical implementation across diverse emergency care settings.

### Comparison with Prior Studies

Several prior studies have explored predictive modeling for outcomes in AF patients in the ED. While these studies have made valuable contributions, many are limited by their methodological design and the granularity of their data. Our findings build upon and extend this prior work, offering significant improvements in both scope and performance.

Liu et al. (2021) [33] developed machine learning models to predict all-cause mortality in 2,037 coronary artery disease patients with AF, with regularized LR achieving the highest AUROC of 0.732 (95% CI: 0.649–0.816). In contrast, our study, using larger MIMIC-IV-ED and MIMIC-IV datasets and incorporating a broader set of tree-based algorithms, found that LightGBM and CatBoost outperformed LR by capturing complex non-linear relationships and feature interactions, providing superior discrimination for ICU transfer and short-term mortality outcomes.

Hong et al. (2025) [28] conducted a large retrospective study on AF patients from the MIMIC-IV database to develop and validate a simplified scoring model. They employed the RF algorithm to predict several outcomes, including ICU transfer and mortality at 3, 7, and 30 days. Their models demonstrated strong discriminative performance in the testing set, with AUROCs of 0.724 for ICU transfer, 0.782 for 3-day mortality, 0.755 for 7-day mortality, and 0.767 for 30-day mortality. In contrast, our study further improves predictive performance and clinical interpretability by employing a hybrid RFECV and LASSO feature selection strategy, reducing the feature set to 19 clinically relevant variables, incorporating SHAP-based explanations, and systematically evaluating multiple machine learning algorithms.

In summary, while prior AF-focused predictive studies have provided valuable foundations for outcome risk stratification in ED patients, our study advances the field by integrating high-resolution MIMIC-IV-ED and MIMIC-IV datasets with a hybrid feature selection strategy, diverse tree-based machine learning algorithms, and SHAP-based interpretability into a unified framework. This approach not only achieves improved predictive performance but also offers clinically meaningful, transparent explanations of individual risk drivers, enhancing the medical interpretability and practical utility of early risk stratification for AF patients in the ED.

## Limitations and Future Work

Despite the robust methodology and improved predictive performance of our LightGBM- and CatBoost-based models, several limitations should be noted. First, the study was conducted retrospectively using the MIMIC-IV-ED and MIMIC-IV databases, which may limit the generalizability of our findings to other EDs or hospital systems. Second, feature extraction relied on structured variables collected at ED triage, capturing only the initial patient assessment and potentially missing subsequent temporal changes or interventions that could influence ICU transfer and short-term mortality risk. Third, although SHAP-based analysis provides interpretable insights, the models have not yet been prospectively validated or tested across multi-center cohorts.

Future work will focus on external validation in diverse ED and hospital settings, the incorporation of longitudinal and time-aware modeling approaches to capture evolving patient trajectories, and the development of clinician-friendly decision support tools that integrate seamlessly with EHR systems, offering real-time, interpretable guidance for early risk stratification of AF patients.

## Conclusion

This study develops and systematically evaluates an interpretable machine learning framework for predicting ICU transfer and short-term mortality in ED patients with AF, leveraging large-scale clinical data from the MIMIC-IV and MIMIC-IV-ED databases. The pipeline incorporated stringent cohort selection, hybrid feature selection, and modular preprocessing to address data heterogeneity and missingness. A clinically coherent subset of predictors—most notably O_2_sat, respiratory rate, heart rate, and systolic blood pressure as acute physiological drivers, alongside age and Charlson Comorbidity Index as markers of frailty and chronic disease burden—was consistently retained, aligning closely with the SHAP-based analyses. This concordance maximized predictive accuracy while ensuring interpretability and clinical plausibility.

Among the models evaluated, LightGBM consistently achieved the highest AUROC for ICU transfer as well as 7- and 30-day mortality, outperforming traditional statistical approaches and other ensemble learners. CatBoost, however, demonstrated superior discrimination for 3-day mortality, highlighting its particular sensitivity to very early physiological deterioration. Together, these complementary strengths underscore the potential value of aligning model choice with the clinical time horizon of interest. LightGBM further exhibited a favorable balance of sensitivity and specificity, a property critical for minimizing missed high-risk patients while limiting false alarms. Feature ablation analysis confirmed the robustness of top-ranked predictors, while SHAP interpretation highlighted the layered contribution of vital signs as acute triggers and frailty-related comorbidities as longer-term modulators of AF-related outcomes.

Clinically, the proposed framework enables early and reliable identification of high-risk AF patients at the point of ED admission using routinely available data. The interpretability afforded by SHAP-based analyses enhances clinician confidence and facilitates integration into real-time decision support systems. Future research should pursue rigorous external validation across multiple centers, prospective evaluation in operational settings, and the incorporation of longitudinal trajectories and multimodal data streams to further improve predictive utility and generalizability.

Taken together, these findings establish a robust and interpretable foundation for AF outcome prediction in emergency care. By delineating a layered risk structure—where acute physiological instability drives immediate outcomes and frailty-related comorbidities modulate longer-term vulnerability—the framework holds promise for guiding timely interventions, optimizing resource allocation, and ultimately improving patient outcomes in aging populations.

## Data Availability

The data underlying the results presented in this study are available from the publicly accessible MIMIC-IV and MIMIC-IV-ED databases, which are hosted on PhysioNet (https://physionet.org/content/mimiciv/ and https://physionet.org/content/mimiciv-ed/). Access to these databases requires registration, completion of the required human subjects training, and agreement to the data use policies. Researchers who meet these criteria can request and obtain the data directly from PhysioNet. All data necessary to replicate the analyses reported in this manuscript are included within the manuscript and its Supporting Information files, and additional details can be provided upon reasonable request.

## Acknowledgments

T.L. independently developed the study concept, designed the research approach, and conducted experiments and data analysis. The manuscript was collaboratively written and critically revised with Z.L. S.C. contributing to the manuscript revision. E.P., K.A., and G.P. provided critical feedback on study design and interpretation of the findings. M.P. oversaw the project, coordinated research activities, and provided strategic guidance throughout. All authors have reviewed and approved the final version of the manuscript.

We also thank the Laboratory for Computational Physiology at the Massachusetts Institute of Technology for their maintenance of the MIMIC-III database.

